# Exploring the Effect of a Supervised Exercise Intervention on Immune Cell Function and Tumour Infiltration in Patients with Breast Cancer Receiving Neoadjuvant Chemotherapy

**DOI:** 10.1101/2025.03.06.25323537

**Authors:** Anna Ubink, Marieke R. ten Tusscher, Hans J. van der Vliet, Joeri A.J. Douma, Tanja D. de Gruijl, Hetty Bontkes, Petra Bonnet, Diede van Ens, Willemijn Hobo, Harry Dolstra, Ellis Barbé, Susanne van der Velde, Catharina Willemien Menke-van der Houven van Oordt, Simone H.C. Havenith, Annemarie Conijn-Mensink, Annette A. van Zweeden, Harm Westdorp, Joannes F.M. Jacobs, Laurien M. Buffart

## Abstract

Pre-clinical studies have shown that exercise can decrease tumour growth through mobilisation, activation, and increased tumour infiltration of natural killer (NK) and CD8^+^ T cells. It is currently unclear whether this can be extrapolated to patients. Therefore, a pilot study was set up to examine the feasibility of obtaining an additional study biopsy and to generate preliminary data on the potential effects of exercise on immune cell function and tumour infiltration. Twenty patients with stage I-III breast cancer receiving neoadjuvant chemotherapy were included (participation rate: 27%). Patients were randomised into the intervention group consisting of a six-week supervised aerobic and resistance exercise program or the control group. Blood samples and tumour biopsies were collected before randomisation and after six weeks of chemotherapy. For 8 of 20 (40%) patients, we were able to obtain and analyse biopsies at diagnosis and six-week follow-up. Upon co-culture of peripheral blood mononuclear cells with K562 tumour cells, the exercise group showed increased expression of the degranulation marker CD107a on NK cells (β=1038.5, 95%CI=56.9; 2020.2, p=0.04), and a trend towards increased tumour cell lysis (β=18.8%, 95%CI=-3.9; 41.5, p=0.10) compared to the control group. In conclusion, the study design was feasible with regard to the participation rate, however, revision is needed with regard to the use of a study-related biopsy. Preliminary evidence was found that exercise during chemotherapy may enhance peripheral NK cell function. Larger studies are needed to extensively study the potential effects of exercise on immune cell function and tumour infiltration in patients with cancer.

## 1. Introduction

Breast cancer is the most common type of cancer in women, with an incidence of approximately 2.3 million new cases worldwide in 2022 [1]. Current treatment often includes chemotherapy [2], which is associated with side effects such as nausea and fatigue [3]. Evidence from randomised controlled trials shows that exercise during cancer treatment is feasible and benefits physical fitness, fatigue, and quality of life [4, 5]. Additionally, observational studies have shown that patients with cancer who are physically active have a lower risk of cancer recurrence and mortality [6, 7]. Yet, the causal effects of exercise on these clinical outcomes and underlying mechanisms of action have not fully been elucidated.

Studies in rodents have shown that exercise can directly reduce tumour growth through mobilisation, activation, and increased tumour infiltration of natural killer (NK) and CD8^+^ T-cells in an epinephrine-dependent manner [8–11]. Similarly, studies in healthy individuals and in patients with breast cancer showed that epinephrine results in mobilisation of predominantly NK cells into the circulation within minutes of initiating exercise [12, 13]. In addition, it has been shown that exercise pre-treatment can enhance peripheral NK cell function in patients with chronic lymphocytic leukaemia [14] and may increase NK cell infiltration into the tumour in patients with prostate cancer [15]. As chemotherapy has been shown to decrease NK cell function [16–20], it is important to understand the immunomodulatory effect of exercise during chemotherapy treatment. Findings from a previous pilot trial in 14 patients with breast and colon cancer suggest that exercise may maintain NK cell function during chemotherapy [20], but the effect on tumour infiltration during chemotherapy has not been described.

To assess the effect of exercise on intra-tumoural immune cell infiltration during chemotherapy, it is necessary to obtain tumour tissue both before and after an exercise intervention period. This can be achieved in the neoadjuvant setting. Depending on cancer stage and molecular subtype, up to 40% of patients with breast cancer achieve pathological complete response after neoadjuvant chemotherapy [21]. Hence, to improve the availability of tumour material for analyses of immune cell infiltration, a biopsy could be obtained before the end of neoadjuvant treatment. However, it is currently unknown whether this is feasible. Therefore, the **S**upervised exercise to **Pr**omote **I**nfiltration of **N**K-cells into the **T**umour (SPRINT) pilot study aimed to examine the feasibility of the exercise trial including additional biopsy sampling in patients with breast cancer receiving neoadjuvant chemotherapy treatment. As a secondary objective, the potential effects of exercise during chemotherapy on immune cell function and tumour infiltration were examined.

## 2. Methods

### 2.1 Design

The SPRINT trial was a multicentre randomised controlled pilot trial comparing a six-week exercise intervention with a usual care control group. The study was approved by the Medical Ethics Committee of the Amsterdam University Medical Centres (EudraCT number NL72539.029.20) and the local ethical boards of all participating hospitals, including Flevo Hospital, Zaans Medical Centre, and Amstelland hospital. The protocol was prospectively registered (Clinicaltrials.gov identifier NCT04704856).

### 2.2 Participants

Patients with early stage (stage I-III) breast cancer who were scheduled for neoadjuvant chemotherapy with four cycles of two- or three-weekly adriamycin and cyclophosphamide, followed by weekly paclitaxel were invited to participate. Patients were excluded when they already participated in structured vigorous aerobic exercise (>60 min per week) and/or resistance exercise (≥two days per week), had comorbidities that contra-indicated moderate-high intensity aerobic or resistance exercise (e.g., severe heart disease), had severe cognitive disorders, used immunosuppressive medication, had a primary or secondary immunodeficiency disorder, were not able to undergo an ultrasound-guided biopsy of the tumour, or were unable to perform basic activities of daily living.

### 2.3 Recruitment and randomisation

Eligible patients were recruited from four Dutch hospitals, i.e. Amsterdam UMC location VUmc, Flevo Hospital, Amstelland Hospital and Zaans Medical Centre. Eligible patients were informed about the study by their treating physician or oncology nurse and asked if they could be contacted by the study coordinator. Patients who chose not to be contacted by the study coordinator or did not want to participate in the study after contact with the study coordinator were asked to provide a reason for non-participation if they were willing to give one. Patients who were willing to participate were invited to the study centre to give written informed consent and undergo baseline measurements. After baseline assessments, patients were stratified for having triple negative breast cancer (yes or no), extent of disease (N- or N+) and frequency of treatment cycles (two-weekly (i.e., dose dense including pegfilgrastim (Neulasta®)) versus three-weekly), and subsequently randomly assigned to one of the two study arms. Randomisation was concealed using the digital CASTOR electronic data capture system that allocated participants to the control or intervention group in a 1:1 ratio with a variable block randomisation. The laboratory technicians and researchers that examined blood and biopsy tissue were blinded to group allocation.

### 2.4 Study arms

Patients in the exercise group followed an exercise program that was shown to be beneficial for physical fitness, fatigue, quality of life and chemotherapy completion rates in patients with breast cancer receiving adjuvant chemotherapy treatment [5]. Exercise sessions took place two times per week for six consecutive weeks, were supervised by an outpatient (oncology) physical therapist close to a patients’ home, and included aerobic and resistance exercises at moderate-to-high intensity. After a short warming-up (5 min), patients performed resistance exercises targeting six large muscle groups for 20 minutes per session, starting with two series of 12 repetitions at 70% of the one-repetition maximum (1RM) and increasing gradually to two series of eight repetitions of 80% 1RM. Aerobic exercises were performed for 30 minutes per session, with an intensity of 50% to 80% of the maximal workload (Wmax) as estimated by the Steep Ramp Test [22]. The 1RM and the Steep Ramp Test were repeated after three weeks to ensure adequate training load. Additionally, patients were advised to be physically active for 30 minutes on three other days per week at Borg level 12-14. Patients from the control group received usual care and were requested to maintain their usual daily physical activities. To limit contamination (increase of exercise in the control group), non-participation and drop-out, the control group was offered the same six-week exercise intervention after the tumour biopsy at six weeks.

### 2.5 Primary outcome measures

The primary outcome was feasibility regarding the proportion of eligible patients willing to participate and the number of tumour biopsy samples that could be adequately examined for immune cell infiltration. We considered the trial feasible if we achieved a participation rate of 20% and were able to successfully examine biopsies for infiltration of CD56^+^ cells (both before and after the intervention period) in 70% of patients. Feasibility was further examined by collecting reasons for non-participation and drop-out rates.

### 2.6 Secondary outcome measures

Physical fitness, physical activity, and health-related quality of life were assessed, and blood samples were drawn at baseline (T0, before start of neoadjuvant chemotherapy) and six weeks after start of chemotherapy (T1, before start of the third chemotherapy cycle in patients with a three-weekly schedule and before start of the fourth cycle in patients with a two-weekly schedule). The baseline measurement of biopsy tissue consisted of tumour tissue collected at diagnosis as part of usual care. After six weeks a study-specific biopsy was obtained. All participants had their last exercise session ranging from one to three days before the T1 measurement. Analyses of blood and tumour biopsies were performed when both T0 and T1 samples were available. Sociodemographics were collected at baseline using a questionnaire. Medical data were retrieved from the medical records.

#### 2.6.1 Physical fitness and activity and quality of life

*Aerobic fitness* was assessed with the Steep Ramp Test (SRT) [22] and the Åstrand-Rhyming test [23], both by using a cycle ergometer. The Steep Ramp Test is a short maximal exercise test, with an increasing work rate of 25 W every 10 seconds until exhaustion. The outcome is the highest achieved wattage and is referred to as Maximum Short Exercise Capacity (MSEC). The peak power output (in watts) can be calculated from the MSEC with the use of a regression equation [24]. The Åstrand-Rhyming test is a submaximal exercise test in which patients are asked to cycle for six minutes on a steady state heart rate (calculated as 180bpm-age, but at least ≥ 120bpm), measured with a V800 Polar system (Polar, Kempele, Finland). The peak oxygen consumption (peakVO_2_) was estimated by applying the work rate and mean heart rate of the fifth and sixth minute to the Åstrand nomogram, with correction for body weight and age [23].

Upper body *muscle strength* was assessed using a hand-held dynamometer (MicroFET) for elbow flexion using a standardized measurement protocol [25]. Lower body muscle strength was assessed with an indirect one-repetition maximum (1-RM) leg press [26].

*Physical activity levels* were examined with the validated Dutch version of the Short Questionnaire to Assess Health-enhancing physical activity (SQUASH) questionnaire [27]. The SQUASH assesses the amount of habitual physical activity during an average week in the past months using questions referring to frequency, intensity and duration of activities [27].

*Health-related Quality of Life (HRQoL)* was assessed with the European Organisation for Research and Treatment of Cancer Quality of Life Questionnaire-Core 30 questionnaire (EORTC QLQ-C30) [28]. HRQoL was evaluated by calculating the summary score, scores range from 0 to 100, with higher scores representing better HRQoL.

#### 2.6.2 Peripheral immune cell profile

The frequency of peripheral immune cell subsets was determined using flow cytometric analysis of fresh blood. Whole blood was incubated with fluorochrome-labelled antibodies (Supplementary Table 1) and after a lyse-no-wash (for quantification of lymphocyte subsets) or lyse-wash procedure using Optilyse B (Beckman Coulter, Brea, CA), events were acquired on a three laser Gallios flow cytometer (Beckman Coulter). Leukocytes and lymphocytes were identified based on side scatter and CD45 positivity and quantified using Trucount beads (BD BioSciences, Becton, NJ). Within lymphocytes, the proportion [%] of B cells (CD3^-^ CD19^+^), NK cells (CD3^-^CD56^+^) and NKT cells (CD3^+^CD56^+^) were identified. The NK cells were further divided into CD56^dim^ (CD56^+^CD16^++^) and CD56^bright^ (CD56^++^CD16^-/+^) subsets. In a second panel, peripheral blood mononuclear cells (PBMCs) were gated based on side scatter and CD45 positivity, from which monocytes and lymphocytes were distinguished based on CD14 expression and forward scatter. From the lymphocytes, the percentage of total T cells (CD3^+^) was determined, which was further divided into CD4^+^ T cells (CD4^+^CD8^-^) and CD8^+^ T cells (CD4^-^ CD8^+^). The CD4^+^ T cells were split into conventional CD4^+^ T cells and regulatory T cells, identifying regulatory T cells based on high CD25 expression. From the conventional CD4^+^ and CD8^+^ T cells, the percentage naïve (T_naïve_) (CD27^+^CD45RA^+^), central memory (T_CM_) (CD27^+^CD45RA^-^), effector memory (T_EM_) (CD27^-^CD45RA^-^), and terminal effector memory CD45RA^+^ (T_EMRA_) (CD27^-^CD45RA^+^) CD4^+^ and CD8^+^ T cells were determined. Gating strategy is shown in Supplementary Figure 1. Analysis was performed using Kaluza software (version 2.1, Beckman Coulter).

#### 2.6.3 Peripheral NK cell phenotype

PBMCs were isolated from fresh blood using density gradient centrifugation with lymphoprep (STEMCELL Technologies, Cologne, Germany). Obtained PBMCs were suspended in freezing solution (75% foetal calf serum / 25% dimethyl sulfoxide) and stored in liquid nitrogen until further analysis. On the day of analysis, cryopreserved PBMCs were thawed and resuspended in brilliant stain buffer (BD Biosciences) containing nanogam (Sanquin blood bank, Nijmegen, The Netherlands). Subsequently, the PBMCs were stained with fluorochrome conjugated antibodies (Supplementary Table 2) and acquired on a CytoFLEX LX flow cytometer (Beckman Coulter). NK cell phenotype was assessed by the expression of NK cell markers on CD56^dim^ NK cells (CD56^+^CD16^+^) and CD56^bright^ NK cells (CD56^++^CD16^-/+^). A reference sample was included at each measurement day to control for inter-assay variation. Manual gating was performed in Kaluza software 2.2. (Beckman Coulter). Gating strategy is shown in Supplementary Figure 2. In addition to manual gating, cluster analysis of NK cells (CD56^+^CD3^-^) was performed with the markers from panel 1 (Supplementary Table 2) using the OMIQ software from Dotmatics (www.omiq.ai). Flow cytometry data was normalised against the reference sample from each batch with CytoNorm. Subsequently, opt-SNE was applied for dimensionality reduction, and 27 metaclusters were identified using FlowSOM analysis.

#### 2.6.4 Peripheral NK cell function

To assess NK cell function, cryopreserved PBMCs were thawed and incubated overnight in absence or presence of 100 U/ml IL-2 (Chiron, Emeryville, CA) and 10 ng/ml IL-15 (Gibco, Waltham, MA), and subsequently co-cultured for four hours with CFSE stained K562 tumour cells at a 10:1 effector to target cell ratio in the presence of 1:50 diluted PE conjugated anti-CD107a (BD Biosciences). During the last three hours of co-culture, the endoplasmic reticulum protein transport inhibitor brefeldin A (10 µg/ml, Sigma, Saint Louis, MO) was added to enhance the detection of intracellular cytokines. After co-culture, cells were stained with fluorochrome-labelled antibodies against surface markers (Supplementary Table 3), and then fixed and permeabilised. Next, permeabilised cells were stained with fluorochrome-labelled antibodies against intracellular markers (Supplementary Table 3), followed by flow cytometric analysis on a BD FACSlyric flow cytometer (BD Biosciences). NK cell (defined as CD45^+^CD3^-^ CD56^+^) function was evaluated by the expression of the early activation marker CD69, the degranulation marker CD107a, and production of granzyme B, and the cytokines IFN-γ and TNF after K562 tumour cell exposure. The median fluorescence intensity (MFI) was taken of the positive cell population. In case of IFN-γ and TNF, the MFI was taken of the total cell population as no clear positive population was present. Target lysis was calculated using the number of CFSE^+^ K562 cells which was quantified using Perfect-Count™ Microspheres (Cytognos, La Serna, Spain), corrected for spontaneous K562 cell death. Analysis was performed using Kaluza software 2.2. (Beckman Coulter). Gating strategy is shown in Supplementary Figure 3.

#### 2.6.5 Immune cell infiltration into the tumour

Tumour biopsies were formalin-fixed and paraffin-embedded, whereafter 4-µm thick tissue sections were obtained. Tissue sections were stained with haematoxylin and eosin (H&E), or immunohistochemically stained for CD56 and granzyme B or CD4 and CD8 using the Ventana Benchmark Ultra immunostainer (Roche/Ventana, Tucson, USA). For details see Supplementary Table 4. Stained slides were digitalised using an Olympus VS200 slide scanner at a magnification of 20x. A pathologist specialised in breast cancer identified areas with tumour tissue within the H&E-stained tissue sections. In these identified tumour areas, quantification of CD56^+^ cells was performed manually as previously described [15]. In brief, CD56^+^ cells were counted within the whole tumour area using Qupath software. Counting was performed by two independent researchers and was repeated when the two measurements differed >20%. In case the two measurements differed <20%, the average number of positive cells was taken. This number of cells was divided by the total tumour area, resulting in CD56^+^ cells cells/mm^2^. Additionally, the CD4/CD8 ratio was determined by counting CD4^+^ and CD8^+^ cells in a random tumour area to a total number of 100 CD4^+^ or CD8^+^ T cells, and then dividing the number of CD4^+^ cells by the number of CD8^+^ cells. In case no tumour tissue was present in the biopsy or when the total tumour area in the biopsy was less than 0.1 mm^2^ [29], immune cell count was not performed.

#### 2.6.6 Statistical analyses

Descriptive data was generated to examine the participation rate and the feasibility of obtaining study-related biopsies. The exercise intervention effects on all outcomes were explored using linear regression analyses in which the intervention was regressed on the post-test value of the outcome (T1), adjusting for the baseline value (T0). The regression coefficients (β), which indicate the between-group differences at T1 corrected for the baseline values, and 95% confidence intervals (95%CI) were reported. Separate models were built for each outcome. Besides between-group differences, within-group differences between T0 and T1 were also examined for the peripheral immune cell profile and NK cell phenotype and function using a paired Wilcoxon signed rank test. Only patients with complete follow-up data were included in all analyses. Additional per-protocol analyses were performed in which patients who received <75% of the exercise sessions were excluded. As this pilot trial is underpowered by design, trends towards statistical significance (p<0.10) of between-group differences were also reported, next to statistically significance differences (p<0.05). Statistical analysis was performed in IBM SPSS (version 29) and R (version 4.1.3) via R studio (version 2022.02.1).

## 3. Results

### 3.1 Feasibility

Of the 73 patients who were eligible for inclusion, 20 (27%) participated in the trial. The characteristics of the included patients are presented in Table 1. Most commonly reported reasons for nonparticipation were practical, emotional or physical in nature (n=16) and the burden of the study-related biopsy (n=14) (Figure 1). We were able to collect biopsy samples at baseline (T0) and six-week follow-up (T1) in 11 out of 20 (55%) patients. Reasons for incomplete biopsy collection were pathological complete response at T1 (n=2), intervention drop-outs due to chemotherapy side-effects (n=3), refusal of second biopsy due to fear and mental well-being (n=2), too low neutrophil values for T1 biopsy (n=1) and disproportionate side effects of T0 biopsy (n=1). For three other patients, immune cell infiltration into the tumour could not be assessed because of insufficient tumour tissue in the biopsy (<0.1 mm^2^). Consequently, the T0 and T1 biopsy could be successfully obtained and analysed for 8 out of 20 (40%) patients. Of these 8 patients, 63% had an ER+/PR+/HER2-tumour and 38% a triple negative tumour, which was respectively 58% and 42% in the other 12 patients. Breast cancer types were also comparable between the patients of whom we successfully analysed the biopsies (75% infiltrating carcinoma not otherwise specified (NOS) and 25% infiltrating lobular carcinoma) compared to the patients in which we did not (67% infiltrating carcinoma NOS and 33% infiltrating lobular carcinoma).

**Figure 1.**
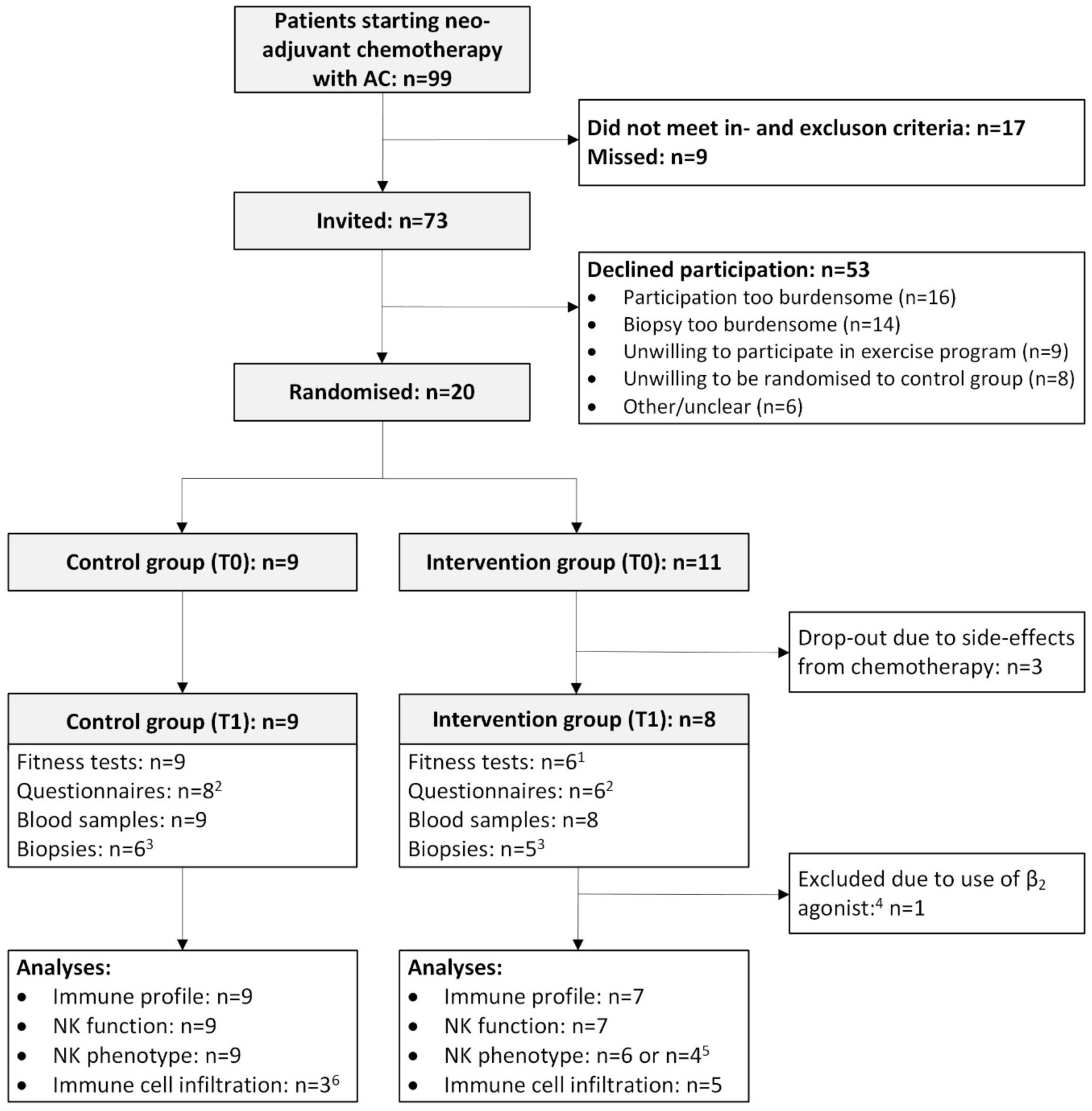
Patient flowchart of the SPRINT pilot study. ^1^Reasons for missing fitness tests were related to low neutrophil values (n=1) or refusal due to poor mental wellbeing (n=1).^2^Three questionnaires were not returned upon request. ^3^Reasons for missing biopsies were pathological complete response (n=2), decline due to fear and poor mental wellbeing (n=2), too low neutrophil values for biopsy (n=1), or disproportionate side effects from diagnostic biopsy (n=1). ^4^One patient was excluded from the immune analyses due to co-medication with salmeterol, which has been shown to affect NK cell phenotype and function [30–32]. ^5^Due to a low amount of isolated PBMCs, in one case only NK cell phenotype panel 1 could be measured and in two cases no NK cell phenotype panels could be measured. ^6^Three biopsies could not be analysed due to limited (<0.1 mm^2^) tumour tissue present in the biopsy. For one additional patient, the CD4/CD8 ratio could not be assessed due to too much necrotic tissue in the biopsy.

**Table 1.**
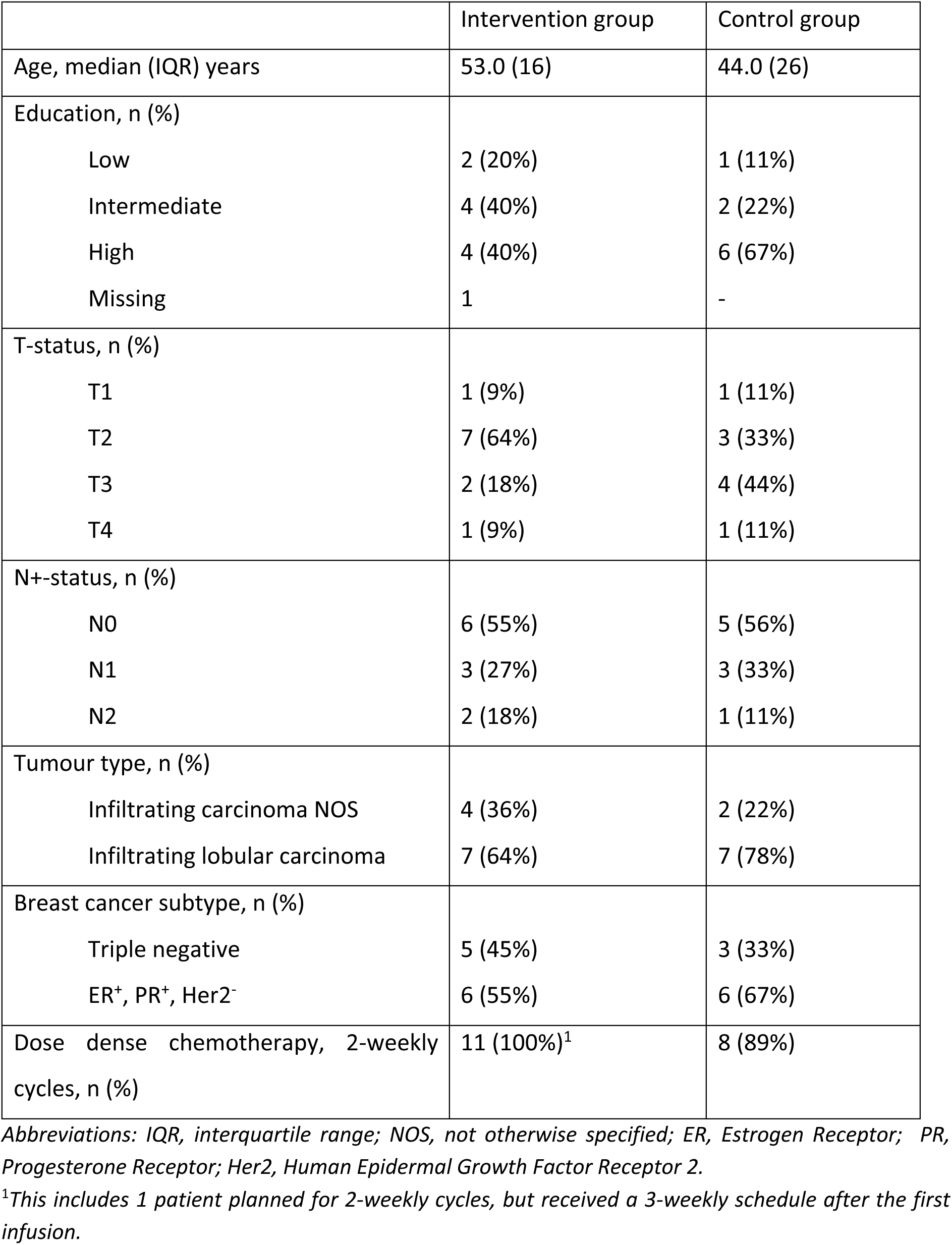
Patient characteristics (n=20)

### 3.2 Physical activity, fitness, and quality of life

The exercise intervention resulted in a trend towards a beneficial effect for elbow flexion strength compared to the control group (β = 23.9 kg, 95%CI = -4.5; 52.4). No significant intervention effects were found for the other outcomes (Table 2).

**Table 2.** Baseline and post-test measurements and between-group differences on physical fitness, physical activity and HRQoL.

| | Intervention group | | Control group | | Between-group difference,<br>$\beta$ (95% CI) |
| --- | --- | --- | --- | --- | --- |
|  | T0 mean (SD) | T1 mean (SD) | T0 mean (SD) | T1 mean (SD) |  |
| MSEC, watts | 261 (44.9) | 251 (41.3) | 253 (42.1) | 231 (51.8) | 11.8 (-19.1; 42.7) |
| PeakVO<sub>2</sub> estimated by the SRT, ml/kg/min | 20.2 (2.4) | 19.6 (2.5) | 20.3 (4.2) | 19.5 (4.5) | 0.3 (-1.3; 1.8) |
| PeakVO<sub>2</sub> estimated by the Åstrand-Rhyming Test, ml/kg/min | 26.9 (0.95) | 23.1 (3.1) | 25.8 (7.2) | 22.7 (9.4) | -0.52 (-8.7; 7.7) |
| Muscle strength |  |  |  |  |  |
| Leg press, kg | 160 (18.8) | 174 (44.4) | 154 (30.6) | 166.7 (40.8) | 2.1 (-43.6; 47.4) |
| Elbow flexion, Nm | 214 (54.5) | 223 (62.9) | 188 (25.7) | 174 (20.2) | 23.9 (-4.5; 52.4) |
| Health-related quality of life (0-100 score) | 75.6 (20.5) | 77.6 (15.4) | 86.2 (13.3) | 78.2 (9.1) | 0.04 (-18.6; 18.7) |
| Self-reported physical activity (total MET-hours/week) | 93.6 (62.5) | 49.3 (57.3) | 89.6 (52.5) | 56.8 (42.3) | -8.9 (-64.6; 46.7) |

*Abbreviations: MSEC, Maximum Short Exercise Capacity; SRT, Steep Ramp Test; MET, Metabolic Equivalent of Task.*

### 3.3 Peripheral immune cell profile

No significant differences between the exercise and control group were found in the absolute counts of leukocytes and lymphocytes or the %monocytes, %B cells, %NK cells, %CD56^dim^ NK cells, %CD56^bright^ NK cells, %NKT cells, %T cells, %regulatory T cells, %CD4^+^ T cells, and %CD8^+^ T cells (Figure 2a-l). In both the exercise and control group, we observed a significant decrease in the absolute lymphocyte count and the %B cells, %CD56^dim^ NK cells, and %CD4^+^ T cells (Figure 2b,d,f,k), as well as a significant increase in the %NKT cells, %T cells, and %CD8^+^ T cells (Figure 2h,i,l) after six weeks of neoadjuvant chemotherapy. The %monocytes and the %CD56^bright^ NK cells significantly increased in the control group, while it remained stable in the exercise group, but this did not result in a significant between-group difference (Figure 2c,g). Results of the per-protocol analysis including patients that followed >75% of the prescribed exercise sessions were comparable (data not shown).

**Figure 2.**
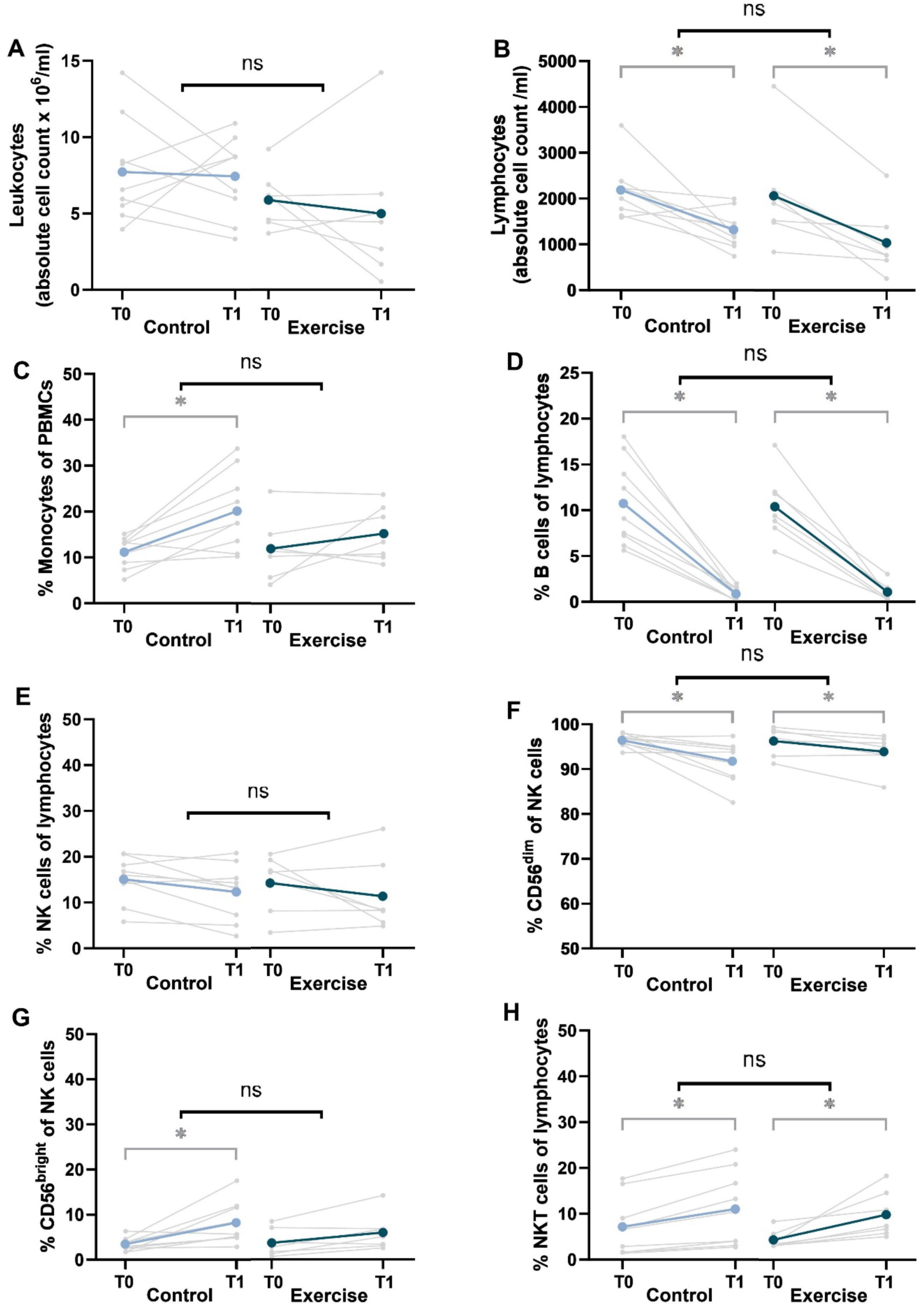
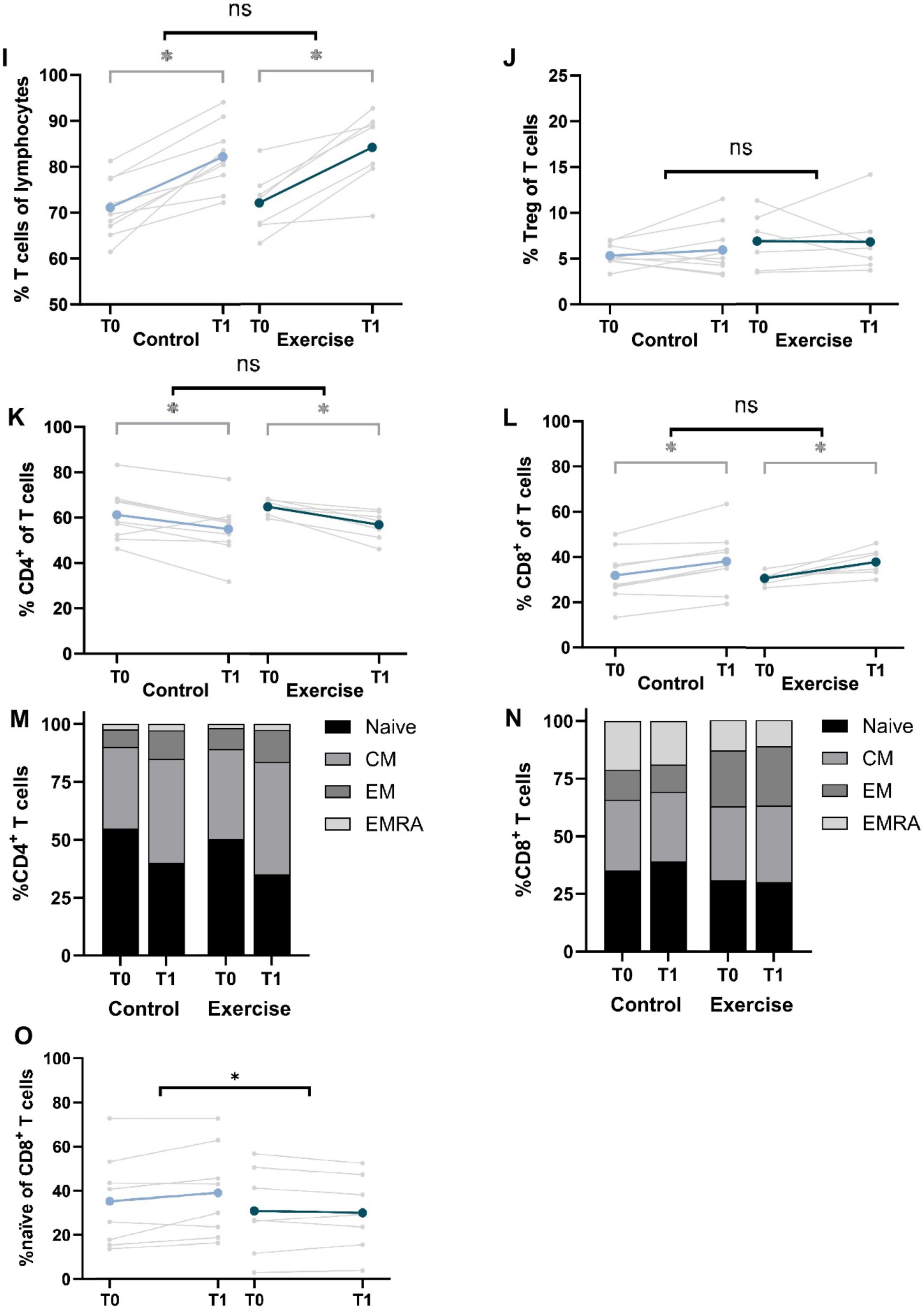
Changes in immune profile from baseline to follow-up in the exercise intervention and control groups. **A)** Absolute counts of leukocytes; **B)** absolute counts of lymphocytes; **C)** %monocytes of peripheral blood mononuclear cells (PBMCs); **D)** %B cells of lymphocytes; **E)** %Natural Killer (NK) cells of lymphocytes; **F)** %CD56^dim^ of NK cells; **G)** %CD56^bright^ of NK cells; **H)** %NKT cells of lymphocytes; **I)** %T cells of lymphocytes; **J)** %regulatory T cells (Treg) of T cells; **K)** %CD4^+^ of T cells; **L)** %CD8^+^ of T cells; **M)** %T_naïve_, %T_CM_, %T_EM_ and %T_EMRA_ of CD4^+^ T cells; **N)** %T_naïve_, %T_CM_, %T_EM_ and %T_EMRA_ of CD8^+^ T cells; and **O)** %T_naïve_ of CD8^+^ cells in peripheral blood of patients with breast cancer from the exercise intervention (n=7) and control group (n=9) before (T0) and after (T1) six weeks of neoadjuvant chemotherapy. Data is presented as individual values and/or mean. Statistically significant differences (p<0.05) are indicated by *. Abbreviations: ns, not significant; CM, central memory; EM, effector memory; EMRA, effector memory CD45RA^+^.

From the CD4^+^ T cells, the %CD4^+^ T_naïve_ cells significantly decreased and the %CD4^+^ T_CM_ and %CD4^+^ T_EM_ cells significantly increased in both groups after chemotherapy with no between-group differences (Figure 2m, Supplementary Table 5). From the CD8^+^ T cells, a significant between-group difference of -5.0% (95%CI = -9.4; -0.6, p = 0.03) was found in the %CD8^+^ T_naïve_ cells, which was maintained in the exercise group while a trend (p = 0.05) towards an increase was seen in the control group after chemotherapy (Figure 2n,o, Supplementary Table 5). However, no between-group difference was seen in the absolute count of CD8^+^ T_naïve_ cells, nor in other CD8^+^ T cell subsets (Supplementary Figure 4). The %CD8^+^ T_EMRA_ cells significantly decreased in the control group while it remained stable in the exercise group, with no significant between-group difference (Figure 2n, Supplementary Table 5). Similar results were seen in the per-protocol analysis (data not shown).

### 3.4 NK cell phenotype

A trend towards a lower expression of the immune checkpoint receptor CD96 (β = -8.2%, 95%CI = -16.8; 0.5, p = 0.06) and the activating receptor NKG2D (β = -3.7%, 95%CI = -7.7; 0.3, p = 0.06) on CD56^dim^ NK cells was observed in the exercise group compared to the control group after 6 weeks of neoadjuvant chemotherapy (Figure 3, Supplementary Table 6). The between-group difference in the %NKG2D^+^ CD56^dim^ NK cells was significant in the per-protocol analysis (β = -5.3%, 95%CI = -9.0; -1.6, p = 0.01). In addition, per-protocol analyses showed a trend towards a lower %NKG2D CD56^bright^ NK cells (β = -3.1%, 95%CI = -6.3; 0.1, p = 0.05) and %CD62L CD56^dim^ NK cells (β = -5.1%, 95%CI = -11.3; 1.1, p = 0.10) in the exercise group compared to the control group after chemotherapy. No between-group differences were found for other NK cell markers (Figure 3, Supplementary Table 6). Cluster analysis of NK cells based on the expression of activating and inhibitory markers also showed a similar distribution of NK cell clusters between exercise and control (Supplementary Figure 5).

**Figure 3.**
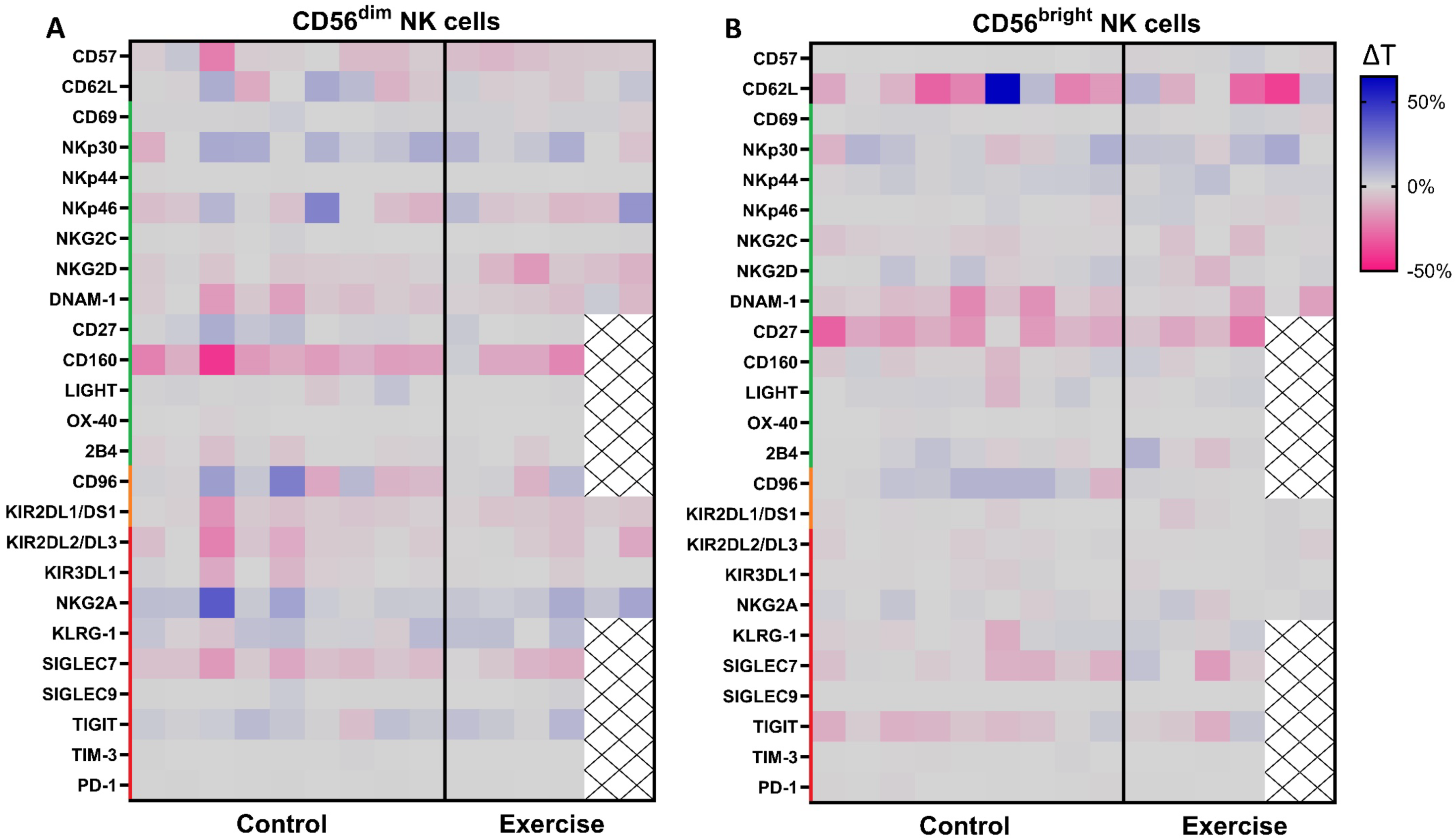
Changes in NK cell phenotype from baseline to follow-up in the exercise intervention and control groups. The absolute change in percentage positive cells between baseline (T0) and after six weeks of neoadjuvant chemotherapy (T1) of NK cell markers on peripheral **A)** CD56^dim^ and **B)** CD56^bright^ NK cells of patients with breast cancer from the exercise intervention (n=4 or 6) and control group (n=9). A trend towards a lower expression of the immune checkpoint receptor CD96 (β = -8.2%, 95%CI = -16.8; 0.5, p = 0.06) and the activating receptor NKG2D (β = -3.7%, 95%CI = -7.7; 0.3, p = 0.06) on CD56^dim^ NK cells was observed in the exercise group compared to the control group after 6 weeks of neoadjuvant chemotherapy. Green = activating, orange = activating/inhibitory, red = inhibitory. Data is presented as individual values (ΔT, %positive NK cells at T1 – %positive NK cells at T0).

### 3.5 NK cell function

The %CD107a^+^ NK cells upon exposure to K562 tumour cells decreased in both the exercise and control group after six weeks of neoadjuvant chemotherapy in the unstimulated condition, with no between-group differences (Figure 4a, Supplementary Table 7). In contrast, the MFI of the degranulation marker CD107a on NK cells was significantly higher in the exercise group compared to the control group (β = 1038.5, 95%CI = 56.9; 2020.2, p = 0.04) (Figure 4b, Supplementary Table 7). The higher expression of CD107a was observed in the cytotoxic CD56^dim^ NK cells, but not in the regulatory CD56^bright^ NK cells (Supplementary Table 7). Corresponding to an enhanced degranulation capacity of NK cells, a trend towards increased lysis of K562 tumour cells was observed in the exercise group compared to the control group (β = 18.8%, 95%CI = 3.9; 41.5, p = 0.10) (Figure 4k). No differences between the groups were found in the percentage positive NK cells and MFI of CD69 (Figure 4c,d), granzyme B (Figure 4e,f), IFN-γ (Figure 4g,h), and TNF (Figure 4i,j). After overnight stimulation of PBMCs with IL-2 and IL-15, no between-group differences in NK cell function were found (Supplementary Table 8). The per-protocol analysis showed similar results (data not shown).

**Figure 4.**
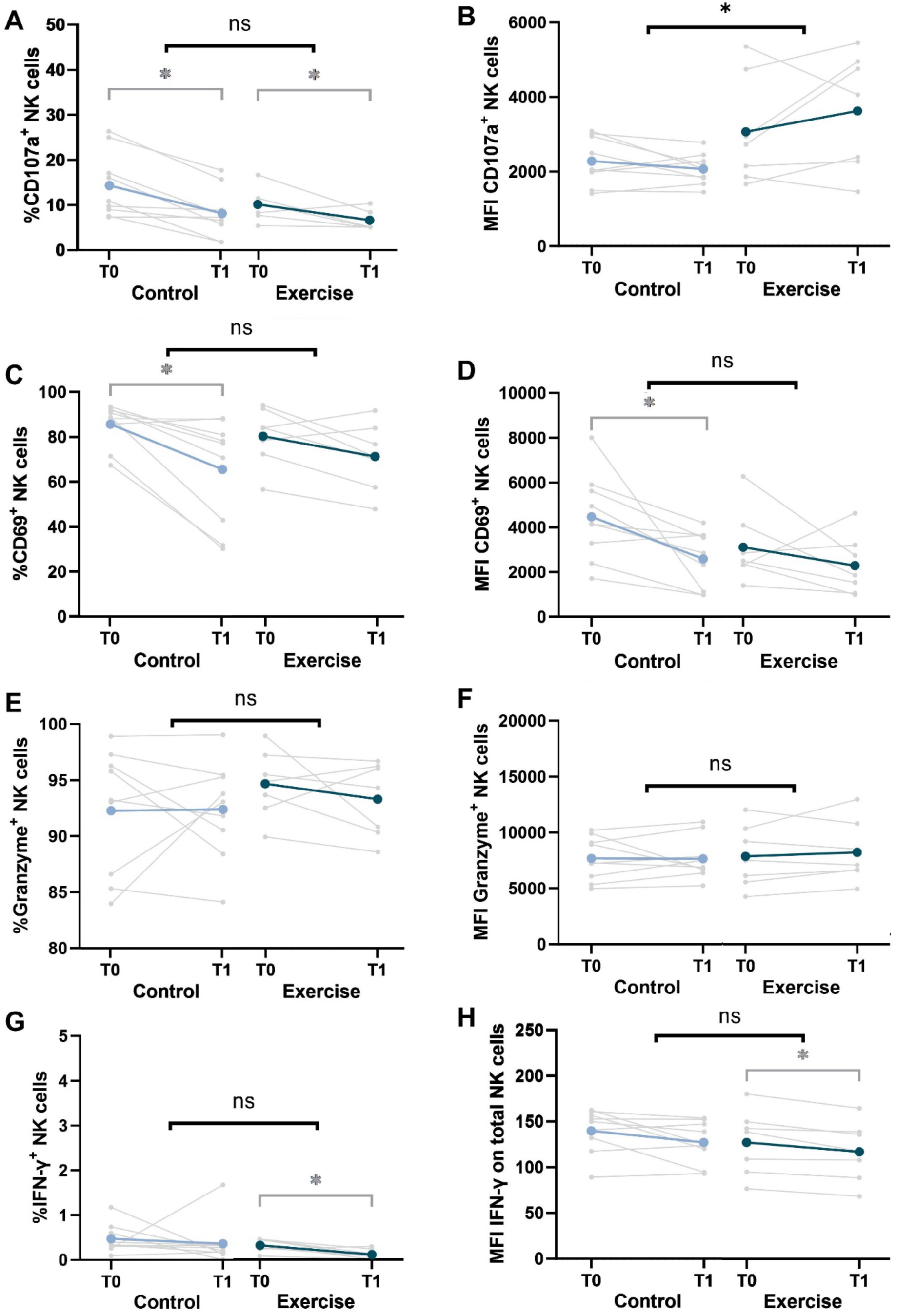
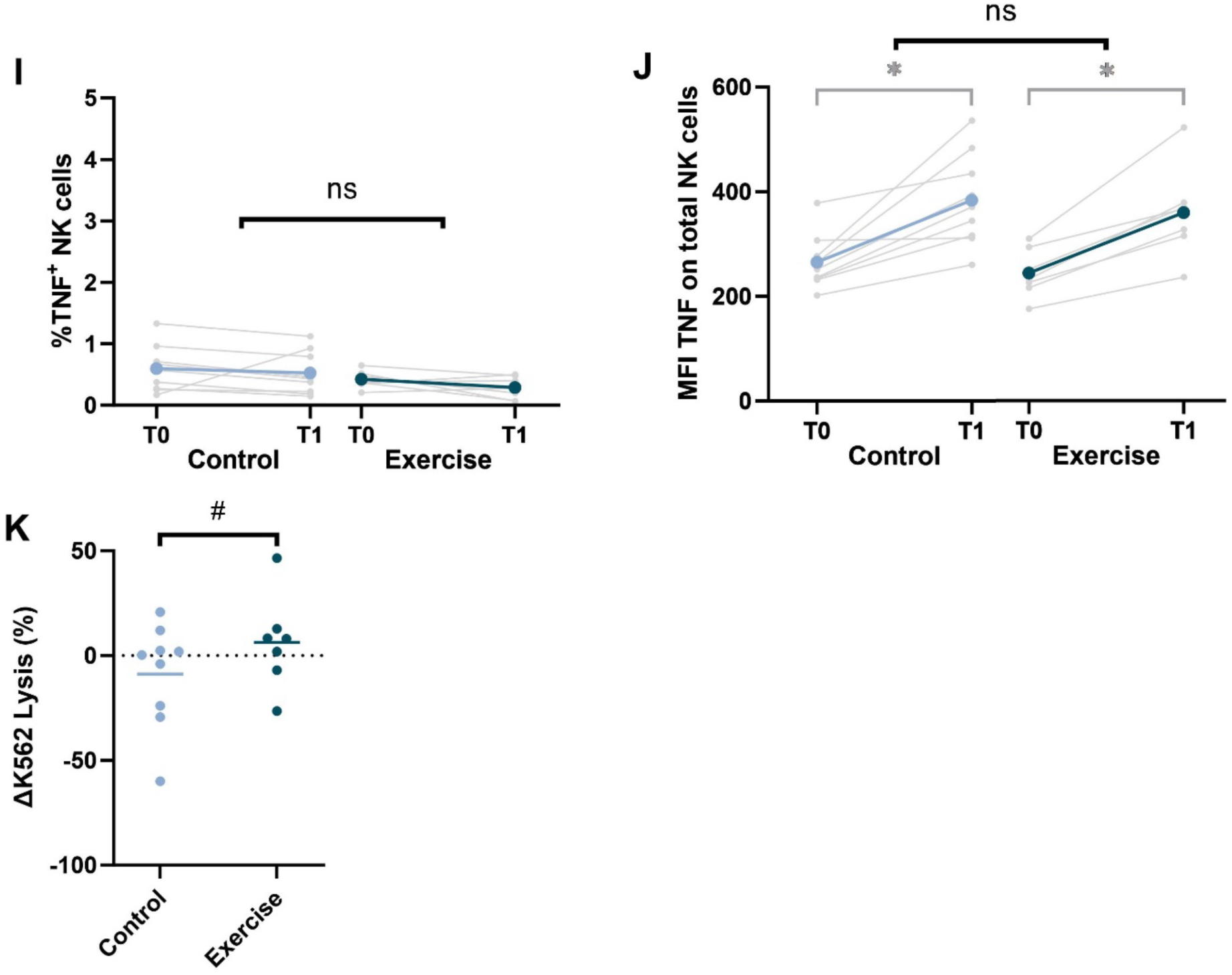
Changes in NK cell function from baseline to follow-up in the exercise intervention and control groups. The percentage positive NK cells and median fluorescence intensity (MFI) of **A, B)** the degranulation marker CD107a; **C, D)** the early activation marker CD69; **E, F)** the cytotoxicity marker granzyme B; **G, H)** the cytokine IFN-γ; and **I, J)** the cytokine TNF after co-culture of unstimulated PBMCs of the exercise intervention (n=7) and control group (n=9) with K562 cells at a 10:1 ratio before (T0) and after (T1) six weeks of neoadjuvant chemotherapy. **K)** The ΔLysis (%) (T1 – T0) of K562 tumour cells after 4h co-culture with unstimulated PBMCs. %Lysis was calculated as the percentage killed K562 cells, corrected for spontaneous K562 cell death. Individual values and mean values are presented. Trends towards statistically significant differences (p<0.10) are indicated by #, and statistically significant differences (p<0.05) are indicated by *.

### 3.6 Immune cell infiltration

The number of CD56^+^ cells / mm^2^ tumour tissue decreased in all three patients of the control group after neoadjuvant chemotherapy, while it remained stable in the exercise group (Figure 5a, Supplementary Figure 6a). In addition, the CD4/CD8 ratio decreased in all five patients from the exercise group, while it increased in one patient and decreased in the other patient from the control group (Figure 5b, Supplementary Figure 6b).

**Figure 5.**
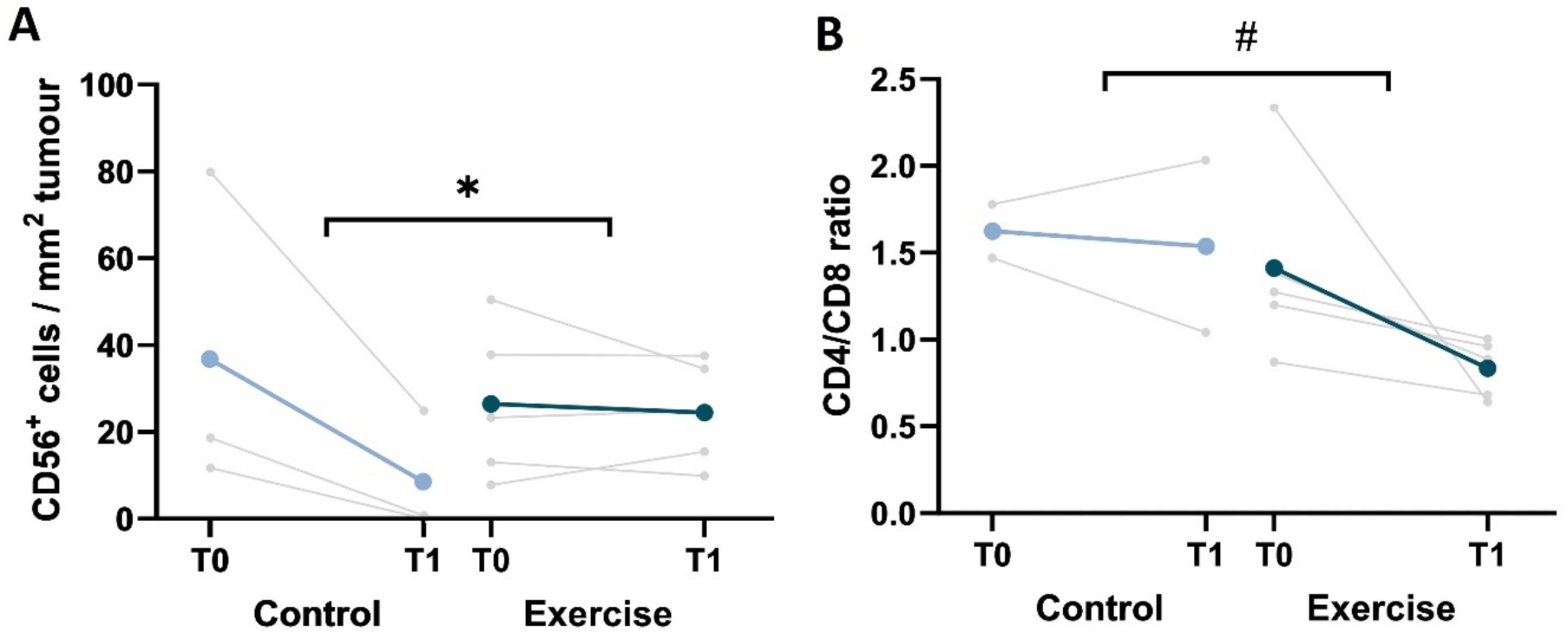
Changes in immune cell infiltration in the tumour from baseline to follow-up in the exercise intervention and control groups. **A)** The number of CD56^+^ cells per mm^2^ tumour tissue and **B)** the CD4/CD8 ratio in the biopsies of the exercise intervention (n=5) and control group (n=3 or n=2) before (T0) and after (T1) six weeks of neoadjuvant chemotherapy. Trends towards statistically significant differences (p<0.10) based on exploratory linear regression analyses are indicated by #, and statistically significant differences (p<0.05) are indicated by *.

## 4. Discussion

This pilot randomised controlled trial examined the feasibility of additional biopsy sampling in an exercise trial, and preliminary effects of a six-week exercise intervention during neoadjuvant chemotherapy on immune cell function and tumour infiltration in patients with breast cancer. We found an acceptable participation rate of 27%, but the feasibility with regard to the study-related biopsy was low due to a limited number of adequately assessable biopsies on both timepoints (40%). Nevertheless, we found preliminary evidence that exercise during chemotherapy may enhance peripheral NK cell function.

The small proportion of patients with evaluable biopsies at both T0 and T1 showed limited feasibility for the present study design. By including a study-related biopsy after six weeks of treatment, we expected to limit missing tumour samples due to pathological complete responses. However, the 15% pathological complete response rate in our study is within the mean pathological complete response rates of 9-33% from studies using resection material in patients with ER^+^/PR^+^/HER2^-^ (range: 5.5-31.3%) and triple negative tumours (range: 20.3-62.2%) [33]. The use of resection material would have prevented missing data due to other biopsy-related problems including insufficient tumour material, or patients being incapable or unwilling to undergo a study-related-biopsy. Hence, regardless of the risk of missing data due to pathological complete responses, the use of resection material in exercise oncology trials may be preferred over study-related biopsy [34] as this likely increases participation rates as well. Next to the use of resection material, it may still be worth considering obtaining biopsies in cancer populations not undergoing this toxic dose-dense chemotherapy treatment. For example, a study among patients with prostate cancer awaiting treatment explored the effects of exercise on NK-cell infiltration in the tumor in the preoperative window for radical prostatectomy and reported that all biopsies could be evaluated on both timepoints [15]. We recommend to actively involve patient representatives in designing such a study.

In contrast to previous studies in patients with breast cancer receiving chemotherapy, we found smaller, and no significant beneficial effects of aerobic and resistance exercise training on physical fitness and HRQoL. While the lack of statistical differences in effect was expected due to the low statistical power, the effect sizes were smaller than expected. This could be related to the shorter intervention duration of 6 weeks, as compared with the recommended duration of 8-12 weeks [35]. Additionally, the intervention dropout rates were somewhat higher compared to previous studies, which may potentially be related to the higher toxicity burden (e.g. anaemia, thrombocytopenia) in patients receiving dose-dense chemotherapy treatment as compared with 3-week cycles [36, 37]. But due to the low number of patients, it is difficult to draw strong conclusions on the increased dropout rates, and whether they differ between patients with different ages, comorbidities or comedication.

Our finding that exercise did not affect the resting levels of main immune cell subsets (e.g., lymphocytes, monocytes, T cells, B cells, and NK cells) in peripheral blood is in line with previous studies in patients with breast cancer [38, 39]. However, these studies did not examine the effect of an exercise intervention on T cell subset composition. In our exploratory analyses we found no difference in the CD4^+^ T cell composition, but we found a lower percentage CD8^+^ T_naïve_ cells in the exercise group compared to the control group. This suggests a higher percentage of antigen experienced CD8^+^ T cells (T_CM_, T_EM_, T_EMRA_), as naïve cells have not yet been primed by antigen presenting cells. This difference may be dependent on IL-15, which is a myokine that is released during exercise and contributes to the homeostatic proliferation and survival of memory CD8^+^ T cells [40, 41]. In addition, lactate and tricarboxylic acid cycle metabolites are produced by skeletal muscle during exercise, which have been shown to enhance the effector profile of CD8^+^ T cells and reduce their expression of the lymphoid-homing marker CD62L [11]. Of note, no between-group differences in the absolute cell counts of the CD8^+^ T cell subsets were found, which questions the potential relevance for the anti-tumour immune response. Therefore, the effect of an exercise intervention on the effector function of peripheral CD8^+^ T cells should be further explored in future studies, which could also take into account their cytotoxic capacity and cytokine production.

The NK cell phenotype was only slightly affected by the exercise intervention, as we found a trend towards a lower %CD96^+^ and %NKG2D^+^ CD56^dim^ NK cells in the exercise group compared to the control group, and no differences in other activating/inhibitory markers. The between-group difference in the %NKG2D^+^ CD56^dim^ NK cells was significant in the per-protocol analyses excluding patients who received <75% of the trainings, which suggests a possible effect of exercise. The signalling function of CD96 remains elusive in humans [42], while NKG2D provides activating signals to NK cells after binding its ligands on tumour cells [43]. Therefore, the decrease in the %NKG2D^+^ CD56^dim^ NK cells contradicts the improved degranulation capacity after exercise. However, the decrease in NKG2D might not directly reduce NK cell function as NK cell activation depends on the sum of signals provided by multiple activating and inhibitory receptors.

Six weeks of exercise did not enhance the percentage of degranulating NK cells upon co-culture with tumour cells in our study both in presence and absence of IL-2/IL-15, which contrasts results from a previous pilot study with a 9–12-week exercise intervention during chemotherapy [20]. However, we did find an increased median expression of CD107a on NK cells in the unstimulated condition, suggesting an enhanced degranulation capacity of NK cells after exercise upon tumour cell exposure. In line with this, a trend towards an increased tumour cell lysis was found. It can be speculated that an enhanced degranulation capacity of NK cells can already be seen after six weeks, while a longer duration of the exercise intervention is needed before NK cells switch towards a degranulating phenotype. The enhanced NK cell function in the exercise group might be dependent on the myokine IL-15, which is released during exercise and is important for NK cell maintenance and activation [40, 44, 45]. When the PBMCs were stimulated with 100 U/ml IL-2 and 10 ng/ml IL-15 overnight before co-culture with K562 tumour cells, no difference in NK cell function was observed between the exercise and the control group. Stimulation with IL-2/IL-15 at these concentrations was perhaps a too strong stimulus to maintain the exercise-induced differences. The IL-15 concentration of 10 ng/ml is regularly used to stimulate NK cells in vitro [1, 3, 4], but is much higher than physiological concentrations after exercise in plasma which are more in the range of 10 pg/ml [5, 6]. Therefore, this may have diminished the exercise-induced differences in NK cell function seen in the unstimulated condition.

The number of CD56^+^ cells / mm^2^ tumour tissue decreased in all three patients of the control group after six weeks of chemotherapy, while it remained stable in most patients of the exercise group. As NK cells and activated or senescent CD8^+^ T cells can express CD56 [46], this might suggest higher levels of NK cells and/or CD8^+^ T cells in the tumours of patients from the exercise group compared to the control group. Similarly, the decreased CD4/CD8 ratio in all patients from the exercise group after neoadjuvant chemotherapy might suggest an increased proportion of cytotoxic CD8^+^ T cells relative to helper or regulatory CD4^+^ T cells in the tumour. However, due to the small sample size this needs extensive examination in larger cohorts. While we found large effect sizes of 1.11 and 1.19 for tumour infiltration of CD56^+^ cells and peripheral NK cell degranulation, respectively, we would recommend future trials to anticipate on a medium effect size of 0.50 or perhaps a large effect size of 0.80. In that case, you would need complete data from 64 or 26 patients per arm (alpha 5%, power 80%), respectively. Future studies should also include CD3 in their immunohistochemical analyses to be able to distinguish NK cells (CD56^+^CD3^-^) from CD56-expressing CD8^+^ T cells (CD56^+^CD3^+^).

An increased tumour infiltration of NK cells and CD8^+^ T cells has been associated with improved prognosis for patients with breast cancer [47, 48]. NK cells can kill tumour cells independent of tumour antigen recognition, making them crucial for targeting MHC-I deficient tumours. In contrast, CD8^+^ T cells can specifically kill tumour cells upon recognition of tumour antigens presented on MHC-I. However, tumour growth may persist despite the presence of these immune cells, particularly in the case of a suppressive tumour microenvironment or when immune cell exhaustion occurs [49]. To enhance the effect of cytotoxic immune cells in the tumour, a pro-inflammatory, “hot” tumour microenvironment is needed. This is characterized by enhanced expression of pro-inflammatory cytokines such as IFN-γ, which can also stimulate the secretion of chemokines (e.g. CXCL9, CXCL10, and CXCL11) to amplify the anti-tumour immune response [49]. While this pro-inflammatory tumour microenvironment is associated with improved prognosis, chronic systemic inflammation is associated with worse prognosis and can contribute to cancer development and progression. For example, high levels of C-reactive protein were significantly associated with worse prognosis in patients with breast cancer [50]. This duality highlights the importance of maintaining a pro-inflammatory immune response within the tumour and an anti-inflammatory systemic profile to achieve better clinical outcomes [51]. As exercise might have the potential to promote infiltration of cytotoxic immune cells into the tumour while reducing systemic inflammation [52], this warrants further investigations.

Strengths of this study include the randomised controlled trial design, which is the gold standard to examine the effect of an intervention [53], and the in-depth analyses of the effect of exercise on peripheral immune cell profile and NK cell phenotype and function. However, this study is limited by the small sample size and the heterogeneous breast cancer population in terms of tumour types. While this is justified for evaluating trial feasibility, our findings of the exploratory analyses of the effects of exercise on immune function and tumour infiltration should be interpreted with caution and studied comprehensively in future randomised controlled trials.

In conclusion, the large number of biopsies that were missing or of insufficient quality to examine immune cell infiltration does not justify the preference of on-treatment biopsies over using resection material. Nevertheless, this study provides some preliminary evidence that exercise during chemotherapy may enhance NK cell function. Larger randomised controlled trials are needed to further examine the effect of physical exercise on immune cell function and tumour infiltration, which may pave the way for using exercise as medicine.

## Data Availability

All data produced in the present study are available upon reasonable request to the authors

## Funding

This study was funded by Stichting Cancer Center Amsterdam, project number 2007041. The contributions of AU and LB were further supported by the NWO-ZonMW Vidi grant (grant number 09150171910017) and the Hypatia Fellowship grant from Radboudumc awarded to LB.

## Declaration of interest

The authors declare that they have no competing interests.

## Acknowledgements

The authors would like to thank all patients for participation in the study. In addition, we would like to thank J.W. Hoekman for his support with the laboratory experiments.

## Author contributions

LB, JD, and HV were involved in the conceptualization of the study design and in the funding acquisition. AU and MT performed data curation and visualisation. AU, MT, LB and JJ performed formal analysis. AU, MT, LB, JJ, HW, and HV were involved in the investigation process. The methodology was set up by AU, MT, LB, JJ, HW, HV, WH, DE, HD, TG, and HB. The project administration was performed by MT and LB. Resources were provided by SV, WM, SH, AC, AZ, HB, PB, and EB. Supervision was performed by LB, HV, JJ and HW. AU, MT, LB, JJ and HW wrote the original draft. All authors reviewed and edited the manuscript.

## Data availability

The datasets used and/or analysed during the current study are available from the corresponding author on reasonable request.

---

**Supplementary Table 1.** Flow cytometry panels immune cell profiling.

| **Antigen** | **Clone** | **Fluorochrome** | **Company** | **Panel** |
| --- | --- | --- | --- | --- |
| CD14 | M5E2 | Pacific Blue | Miltenyi Biotec | 2 |
| CD16 | B73.1 | FITC | Biolegend | 1 |
| CD19 | J3-119 | ECD | Beckman Coulter | 1 |
| CD25 | 2A3 | PE | BD Biosciences | 2 |
| CD27 | 1A4CD27 | PC7 | Beckman Coulter | 2 |
| CD3 | BW264/56 | VioGreen | Miltenyi Biotec | 1 |
| CD4 | SK3 | APC-H7 | BD Biosciences | 2 |
| CD45 | 2D1 | PerCP | BD Biosciences | 1,2 |
| CD45RA | 2H4 | ECD | Beckman Coulter | 2 |
| CD56 | HCD56 | BV421 | Biolegend | 1 |
| CD8 | B9.11 | APC-AF700 | Beckman Coulter | 2 |

**Supplementary Table 2.** Flow cytometry panels NK cell phenotype.

| Antigen | Clone | Fluorochrome | Company | Panel |
| --- | --- | --- | --- | --- |
| CD134 (OX40) | Ber-ACT35 (ACT35) | BV786 | BD Biosciences | 2 |
| CD158a,h (KIR2DL1/DS1) | HP-MA4 | FITC | BioLegend | 1 |
| CD158b (KIR2DL2/DL3) | CH-L | BV510 | BD Biosciences | 1 |
| CD158e1 (KIR3DL1) | DX9 | AF700 | BioLegend | 1 |
| CD159a (NKG2A) | Z199 | APC | Beckman Coulter | 1 |
| CD159c (NKG2C) | 134591 | BV711 | BD Biosciences | 1 |
| CD16 | 3G8 | BUV496 | BD Biosciences | 1,2 |
| CD160 | BY55 | PE-Cy7 | BioLegend | 2 |
| CD226 (DNAM-1) 11A8 | 11A8 | BV510 | BioLegend | 2 |
| CD226 (DNAM-1) 11A8 | 11A8 | PE | BioLegend | 1 |
| CD244 (2B4) | C1.7 | PerCP-Cy5.5 | BioLegend | 2 |
| CD258 (LIGHT) | 7-3 (7) | PE | eBioscience | 2 |
| CD27 | O323 | PE-Cy5 | eBioscience | 2 |
| CD279 (PD-1) | EH12.2H7 | BV711 | BioLegend | 2 |
| CD3 | UCHT1 | BUV737 | BD Biosciences | 2 |
| CD3 | UCHT1 | BV605 | BioLegend | 1 |
| CD314 (NKG2D) | 1D11 | BV785 | BioLegend | 1 |
| CD335 (NKp46) | 9E2 | PE-Dazzle594 | BioLegend | 1 |
| CD336 (NKp44) | P44-8 | PE-Cy7 | BioLegend | 1 |
| CD337 (NKp30) | Z25 | PE-Cy5 | Beckman Coulter | 1 |
| CD366 (TIM-3) | F38-2E2 | BV605 | BioLegend | 2 |
| CD45 | HI30 | BUV395 | BD Biosciences | 1,2 |
| CD56 | HCD56 | BV421 | BioLegend | 1,2 |
| CD57 | QA17A04 | PerCP-Cy5.5 | BioLegend | 1 |
| CD62L | DREG-56 | APC-Fire750 | BioLegend | 1 |
| CD69 | FN50 | BUV737 | BD Biosciences | 1 |
| CD96 | NK92.39 | PE-Dazzle594 | BioLegend | 2 |
| KLRG-1 | REA261 | FITC | Miltenyi Biotec | 2 |
| Live/dead | - | ViaKrome808 | Beckman Coulter | 1,2 |
| SIGLEC7 | 6-434 | APC-Fire750 | BioLegend | 2 |
| SIGLEC9 | 191240 | AF700 | R&D systems | 2 |
| TIGIT | 741182 | APC | R&D systems | 2 |

**Supplementary Table 3.** Flow cytometry panel NK cell function.

| Fluorochrome | Antigen | Clone | Company |
| --- | --- | --- | --- |
| APC | CD69 | FN50 | Biolegend |
| BV421 | TNF* | MAb11 | Biolegend |
| BV605 | CD3 | HIT3alpha | BD Biosciences |
| BV711 | CD56 | NCAM16.2 | BD Biosciences |
| E780 | Fixable viability dye | - | eBioscience |
| KO | CD45 | J33 | Beckman Coulter |
| PE-Cy7 | IFN-γ* | 4S.B3 | Invitrogen |
| PerCP-Cy5.5 | Granzyme B* | QA16A02 | Biolegend |

\* *intracellular*

**Supplementary Table 4.** Immunohistochemistry protocols.

| Antigen | Clone | Company | Ventana Ultra Detection method* |
| --- | --- | --- | --- |
| CD4 | SP35 | ABCAM | Optiview detection kit (incl. DAB and CuSO<sub>4</sub>) |
| CD8 | 8/144B | DAKO | Ultraview Alkaline Phosphatase Red |
| Granzyme B | GB7 | Monosan | Optiview detection kit (incl. DAB and CuSO<sub>4</sub>) |
| CD056 | MRQ-42 | Cellmarque | Ultraview Alkaline Phosphatase Red |

\*Tissue sections were pre-treated with CC1 for 24 minutes at 100°C for antigen retrieval, whereafter the primary antibody against granzyme B (Monosan, Uden, The Netherlands) or CD4 (Abcam, Cambridge, UK) was used in combination with the Optiview detection kit including DAB and CuSO<sub>4</sub> (Roche, Basel, Switzerland) for detection and visualisation. Next, slides were treated again with CC1, and incubated with the primary antibody against CD56 (Cellmarque, Rocklin, USA) or CD8 (Dako, Glostrup, Denmark) which was detected with the Ultraview Alkaline Phosphatase Red. After immunohistochemical staining the sections were washed with EZ prep and soap, water, and dehydrated with baths of ethanol and cleared with xylene. All sections were mounted with Tissue Tek<sup>®</sup> cover slipping film (Sakura Finetek Europe B.V., Alphen aan den Rijn, The Netherlands).

**Supplementary Table 5.** Absolute counts or percentages (%) of immune cell subsets in peripheral blood of the exercise intervention (n=7) and control group (n=9) before (T0) and after (T1) 6 weeks of chemotherapy. The regression coefficients (β) indicate the between-group differences corrected for baseline values, as determined by linear regression analysis.

| Variable | N | T0 | T1 | Between-Group change |  |
| --- | --- | --- | --- | --- | --- |
|  |  | Mean (SD) | Mean (SD) | β (95% CI) | p value |
| **<u>Leukocytes (absolute cell count, x 10<sup>6</sup>/ml)</u>** |  |  |  |  |  |
| Exercise | 7 | 5.9 (1.9) | 5.0 (4.5) | -1.8 (-5.9; 2.3) | 0.36 |
| Control | 9 | 7.7 (3.4) | 7.4 (2.6) |  |  |
| **<u>Lymphocytes (absolute cell count /ml)</u>** |  |  |  |  |  |
| Exercise | 7 | 2062.7 (1146.6) | 1038.9 (726.8)* | -236.3 (-782.7; 310.1) | 0.37 |
| Control | 9 | 2184.0 (601.8) | 1317.4 (417.5)* |  |  |
| **<u>% Monocytes of PBMCs</u>** |  |  |  |  |  |
| Exercise | 7 | 11.9 (6.7) | 15.2 (5.9) | -5.4 (-13.1; 2.4) | 0.16 |
| Control | 9 | 11.2 (3.4) | 20.1 (8.5)* |  |  |
| **<u>% B cells of CD45<sup>+</sup> cells</u>** |  |  |  |  |  |
| Exercise | 7 | 10.4 (3.7) | 1.1 (1.0)* | 0.2 (-0.6; 1.1) | 0.55 |
| Control | 9 | 10.7 (4.7) | 0.9 (0.7)* |  |  |
| **<u>% NK cells of lymphocytes</u>** |  |  |  |  |  |
| Exercise | 7 | 14.2 (6.2) | 11.3 (7.8) | -0.3 (-6.1; 5.6) | 0.92 |
| Control | 9 | 15.1 (5.0) | 12.3 (6.2) |  |  |
| **<u>% CD56<sup>dim</sup> of NK cells</u>** |  |  |  |  |  |
| Exercise | 7 | 96.3 (3.1) | 93.9 (3.9)* | 2.3 (-1.9; 6.5) | 0.25 |
| Control | 9 | 96.4 (1.4) | 91.8 (4.7)* |  |  |
| **<u>% CD56<sup>bright</sup> of NK cells</u>** |  |  |  |  |  |
| Exercise | 7 | 3.7 (3.0) | 6.0 (4.0) | -2.5 (-6.8; 1.8) | 0.24 |
| Control | 9 | 3.4 (1.5) | 8.2 (4.7)* |  |  |
| **<u>% NKT cells of lymphocytes</u>** |  |  |  |  |  |
| Exercise | 7 | 4.4 (2.0) | 9.9 (5.0)* | 2.3 (-1.7; 6.4) | 0.24 |
| Control | 9 | 7.2 (6.3) | 11.0 (8.1)* |  |  |
| | | Mean (SD) | Mean (SD) | $\beta$ (95% CI) | p value |
| **<u>% T cells of CD45<sup>+</sup> cells</u>** |  |  |  |  |  |
| Exercise | 7 | 72.0 (6.7) | 84.1 (8.2)* | 1.2 (-5.2; 7.7) | 0.69 |
| Control | 9 | 71.1 (6.5) | 82.2 (7.3)* |  |  |
| **<u>% conventional CD4<sup>+</sup> T cells of T cells</u>** |  |  |  |  |  |
| Exercise | 7 | 65.0 (3.3) | 57.0 (6.4)* | -1.4 (-8.1; 5.3) | 0.66 |
| Control | 9 | 61.2 (11.5) | 54.9 (12.1)* |  |  |
| ***% naïve of conventional CD4<sup>+</sup> T cells*** |  |  |  |  |  |
| Exercise | 7 | 50.2 (10.5) | 35.0 (12.6)* | 0.9 (-8.7; 10.5) | 0.84 |
| Control | 9 | 54.8 (11.2) | 40.1 (18.7)* |  |  |
| ***% central memory of conventional CD4<sup>+</sup> T cells*** |  |  |  |  |  |
| Exercise | 7 | 39.0 (12.1) | 48.6 (14.9)* | -0.2 (-7.3; 7.0) | 0.96 |
| Control | 9 | 35.3 (9.3) | 44.9 (10.9)* |  |  |
| ***% effector memory of conventional CD4<sup>+</sup> T cells*** |  |  |  |  |  |
| Exercise | 7 | 9.1 (7.5) | 13.8 (11.8)* | -0.9 (-2.5; 0.6) | 0.22 |
| Control | 9 | 7.6 (4.6) | 12.3 (8.5)* |  |  |
| ***% EMRA of conventional CD4<sup>+</sup> T cells*** |  |  |  |  |  |
| Exercise | 7 | 1.8 (1.6) | 2.6 (2.8) | 0.5 (-0.9; 2.0) | 0.43 |
| Control | 9 | 2.3 (3.0) | 2.7 (4.2) |  |  |
| **<u>% Regulatory CD4<sup>+</sup> T cells of T cells</u>** |  |  |  |  |  |
| Exercise | 7 | 6.9 (2.9) | 6.8 (3.5) | -0.5 (-3.6; 2.7) | 0.75 |
| Control | 9 | 5.3 (1.2) | 6.0 (2.8) |  |  |
| **<u>% CD8<sup>+</sup> T cells of T cells</u>** |  |  |  |  |  |
| Exercise | 7 | 30.7 (2.7) | 38.0 (5.6)* | 1.1 (-4.0; 6.2) | 0.64 |
| Control | 9 | 31.9 (11.4) | 38.1 (13.1)* |  |  |
| | | Mean (SD) | Mean (SD) | $\beta$ (95% CI) | p value |
| **% naïve of CD8<sup>+</sup> T cells** |  |  |  |  |  |
| Exercise | 7 | 30.8 (19.8) | 30.0 (17.3) | -5.0 (-9.4; -0.6) | **0.03** |
| Control | 9 | 35.2 (19.6) | 39.0 (19.4)<sup>a</sup> |  |  |
| **% central memory of CD8<sup>+</sup> T cells** |  |  |  |  |  |
| Exercise | 7 | 32.0 (13.1) | 33.0 (14.6) | 1.6 (-4.3; 7.5) | 0.57 |
| Control | 9 | 30.6 (11.3) | 30.2 (10.2) |  |  |
| **% effector memory of CD8<sup>+</sup> T cells** |  |  |  |  |  |
| Exercise | 7 | 24.0 (22.0) | 25.7 (21.9) | 3.3 (-1.4; 8.0) | 0.15 |
| Control | 9 | 13.0 (7.6) | 11.8 (7.3) |  |  |
| **% EMRA of CD8<sup>+</sup> T cells** |  |  |  |  |  |
| Exercise | 7 | 13.2 (9.9) | 11.2 (8.5) | -0.7 (-5.1; 3.7) | 0.75 |
| Control | 9 | 21.2 (24.3) | 18.9 (22.0)* |  |  |

\*Significant within-group difference between T0 and T1 based on Wilcoxon signed rank ( $p < 0.05$ ). <sup>a</sup> $p = 0.05$ .

**Supplementary Table 6.** The %positive CD56^dim^ and CD56^bright^ NK cells for NK cell markers of the exercise intervention and control group before (T0) and after (T1) 6 weeks of chemotherapy. The regression coefficients (β) indicate the between-group differences corrected for baseline values, as determined by linear regression analysis.

| Variable | N | T0 | T1 | Between-group change |  |
| --- | --- | --- | --- | --- | --- |
| | | Mean (SD) | Mean (SD) | $\beta$ (95% CI) | p value |
| **%NKp30<sup>+</sup> CD56<sup>dim</sup> NK cells** |  |  |  |  |  |
| Exercise | 6 | 45.1 (10.0) | 48.8 (9.5) | -1.0(-9.6; 7.7) | 0.81 |
| Control | 9 | 41.3 (9.3) | 46.4 (11.7) |  |  |
| **%NKp44<sup>+</sup> CD56<sup>dim</sup> NK cells** |  |  |  |  |  |
| Exercise | 6 | 0.1 (0.1) | 0.2 (0.1) | -0.0 (-0.2; 0.1) | 0.54 |
| Control | 9 | 0.2 (0.4) | 0.2 (0.2) |  |  |
| **%NKp46<sup>+</sup> CD56<sup>dim</sup> NK cells** |  |  |  |  |  |
| Exercise | 6 | 76.7 (10.1) | 77.8 (1.0) | 1.0 (-6.4; 8.4) | 0.78 |
| Control | 9 | 76.3 (8.8) | 76.7 (8.0) |  |  |
| **%NKG2C<sup>+</sup> CD56<sup>dim</sup> NK cells** |  |  |  |  |  |
| Exercise | 6 | 1.2 (0.8) | 1.4 (1.0) | 0.2 (-0.6; 1.0) | 0.54 |
| Control | 9 | 1.9 (1.5) | 1.9 (1.7) |  |  |
| **%NKG2D<sup>+</sup> CD56<sup>dim</sup> NK cells** |  |  |  |  |  |
| Exercise | 6 | 91.2 (5.8) | 84.5 (10.6) | -3.7 (-7.7; 0.3) | **0.06** |
| Control | 9 | 94.2 (6.3) | 92.0(6.5)* |  |  |
| **%NKG2A<sup>+</sup> CD56<sup>dim</sup> NK cells** |  |  |  |  |  |
| Exercise | 6 | 45.2 (14.7) | 52.2 (13.3)* | -1.8 (-11.3; 7.7) | 0.69 |
| Control | 9 | 45.2 (15.1) | 54.1 (12.3)* |  |  |
| **%KIR2DL1/DS1<sup>+</sup> CD56<sup>dim</sup> NK cells** |  |  |  |  |  |
| Exercise | 6 | 26.1 (12.0) | 22.4 (12.5)* | 1.1 (-3.6; 5.9) | 0.61 |
| Control | 9 | 22.6 (6.5) | 18.1 (5.9)* |  |  |
| **%KIR2DL2/DL3<sup>+</sup> CD56<sup>dim</sup> NK cells** |  |  |  |  |  |
| Exercise | 6 | 28.4 (12.9) | 24.9 (10.4) | 0.1 (-5.8; 6.0) | 0.98 |
| Control | 9 | 34.4 (7.6) | 28.6 (5.6)* |  |  |
| **%KIR3DL1<sup>+</sup> CD56<sup>dim</sup> NK cells** |  |  |  |  |  |
| Exercise | 6 | 14.9 (17.6) | 14.2 (17.3) | 1.6 (-2.3; 5.4) | 0.40 |
| Control | 9 | 19.6 (12.3) | 17.0 (11.5)* |  |  |
| **%CD57<sup>+</sup> CD56<sup>dim</sup> NK cells** |  |  |  |  |  |
| Exercise | 6 | 23.8 (9.5) | 18.9 (7.9)* | -1.1 (-7.4; 5.3) | 0.73 |
| Control | 9 | 27.2 (13.4) | 22.6 (12.1) |  |  |
| | | Mean (SD) | Mean (SD) | $\beta$ (95% CI) | p value |
| **%CD62L<sup>+</sup> CD56<sup>dim</sup> NK cells** |  |  |  |  |  |
| Exercise | 6 | 8.4 (3.8) | 8.4 (2.3) | -4.4 (-10.1; 1.4) | 0.12 |
| Control | 9 | 12.6 (7.5) | 13.5 (5.6) |  |  |
| **%CD69<sup>+</sup> CD56<sup>dim</sup> NK cells** |  |  |  |  |  |
| Exercise | 6 | 3.1 (2.8) | 3.4 (1.6) | -0.3 (-1.7; 1.2) | 0.69 |
| Control | 9 | 1.9 (0.9) | 2.9 (1.6)* |  |  |
| **%DNAM-1<sup>+</sup> CD56<sup>dim</sup> NK cells** |  |  |  |  |  |
| Exercise | 6 | 94.1 (1.9) | 90.9 (3.0) | 3.0 (-1.6; 7.6) | 0.18 |
| Control | 9 | 94.7 (3.7) | 88.9 (8.3)* |  |  |
| **%2B4<sup>+</sup> CD56<sup>dim</sup> NK cells** |  |  |  |  |  |
| Exercise | 4 | 96.9 (1.9) | 96.3 (1.6) | 1.2 (-1.5; 4.0) | 0.34 |
| Control | 9 | 96.2 (2.0) | 94.5 (2.8)* |  |  |
| **%LIGHT<sup>+</sup> CD56<sup>dim</sup> NK cells** |  |  |  |  |  |
| Exercise | 4 | 0.5 (0.31) | 0.6 (0.19) | -1.1 (-3.38; 1.23) | 0.32 |
| Control | 9 | 1.0 (1.23) | 1.7 (1.86) |  |  |
| **%OX40<sup>+</sup> CD56<sup>dim</sup> NK cells** |  |  |  |  |  |
| Exercise | 4 | 0.3 (0.2) | 0.3 (0.2) | -0.0 (-0.6; 0.6) | 0.93 |
| Control | 9 | 0.6 (0.8) | 0.5 (0.7) |  |  |
| **%CD160<sup>+</sup> CD56<sup>dim</sup> NK cells** |  |  |  |  |  |
| Exercise | 4 | 31.7 (4.3) | 21.4 (8.5) | -0.5 (-15.3; 14.2) | 0.94 |
| Control | 9 | 45.0 (10.3) | 28.2 (9.8)* |  |  |
| **%KLRG-1<sup>+</sup> CD56<sup>dim</sup> NK cells** |  |  |  |  |  |
| Exercise | 4 | 53.6 (15.9) | 58.7 (13.9) | 3.1 (-2.1; 8.3) | 0.22 |
| Control | 9 | 51.2 (21.9) | 53.5 (20.2) |  |  |
| **%SIGLEC7<sup>+</sup> CD56<sup>dim</sup> NK cells** |  |  |  |  |  |
| Exercise | 4 | 84.3 (6.2) | 78.7 (10.3) | 1.9 (-4.3; 8.2) | 0.51 |
| Control | 9 | 90.0 (6.3) | 83.0 (6.8)* |  |  |
| **%SIGLEC9<sup>+</sup> CD56<sup>dim</sup> NK cells** |  |  |  |  |  |
| Exercise | 4 | 0.4 (0.4) | 0.8 (0.8) | -0.2 (-1.4; 1.0) | 0.72 |
| Control | 9 | 0.5 (0.4) | 1.1 (0.9)* |  |  |
| | | Mean (SD) | Mean (SD) | $\beta$ (95% CI) | p value |
| **%TIGIT<sup>+</sup> CD56<sup>dim</sup> NK cells** |  |  |  |  |  |
| Exercise | 4 | 46.8 (15.81) | 51.6 (13.3) | 1.4 (-3.3; 6.1) | 0.53 |
| Control | 9 | 51.6 (10.30) | 54.3 (9.6) |  |  |
| **%CD96<sup>+</sup> CD56<sup>dim</sup> NK cells** |  |  |  |  |  |
| Exercise | 4 | 26.2 (9.8) | 26.5 (4.1) | -8.2 (-16.8; 0.5) | **0.06** |
| Control | 9 | 32.2 (11.0) | 35.3 (6.6) |  |  |
| **%TIM-3<sup>+</sup> CD56<sup>dim</sup> NK cells** |  |  |  |  |  |
| Exercise | 4 | 0.2 (0.2) | 0.1 (0.1) | -0.2 (-0.5; 0.2) | 0.29 |
| Control | 9 | 0.3 (0.2) | 0.4 (0.4) |  |  |
| **%PD-1<sup>+</sup> CD56<sup>dim</sup> NK cells** |  |  |  |  |  |
| Exercise | 4 | 0.2 (0.1) | 0.3 (0.2) | 0.0 (-0.3; 0.3) | 0.98 |
| Control | 9 | 0.4 (0.2) | 0.5 (0.3) |  |  |
| **%CD27<sup>+</sup> CD56<sup>dim</sup> NK cells** |  |  |  |  |  |
| Exercise | 4 | 4.5 (1.0) | 5.6 (1.9) | -2.5 (-7.2; 2.3) | 0.28 |
| Control | 9 | 4.8 (1.5) | 8.3 (3.9)* |  |  |
| **%NKp30<sup>+</sup> CD56<sup>bright</sup> NK cells** |  |  |  |  |  |
| Exercise | 6 | 18.3 (5.8) | 22.8 (5.0) | 2.4 (-4.2; 9.0) | 0.45 |
| Control | 9 | 19.8 (7.6) | 21.3 (7.9) |  |  |
| **%NKp44<sup>+</sup> CD56<sup>bright</sup> NK cells** |  |  |  |  |  |
| Exercise | 6 | 3.7 (1.5) | 6.2 (3.3)* | -0.0 (-2.7; 2.6) | 0.97 |
| Control | 9 | 2.2 (1.0) | 4.4 (1.5)* |  |  |
| **%NKp46<sup>+</sup> CD56<sup>bright</sup> NK cells** |  |  |  |  |  |
| Exercise | 6 | 97.0 (2.2) | 97.2 (1.8) | -0.5 (-2.1; 1.1) | 0.53 |
| Control | 9 | 98.3 (0.7) | 98.1 (0.9) |  |  |
| **%NKG2C<sup>+</sup> CD56<sup>bright</sup> NK cells** |  |  |  |  |  |
| Exercise | 6 | 4.5 (5.3) | 2.4 (2.4) | -0.2 (-1.4; 1.0) | 0.71 |
| Control | 9 | 4.9 (1.8) | 2.8 (1.6)* |  |  |
| **%NKG2D<sup>+</sup> CD56<sup>bright</sup> NK cells** |  |  |  |  |  |
| Exercise | 6 | 91.1 (9.8) | 89.9 (9.4) | -2.5 (-5.6; 0.7) | 0.11 |
| Control | 9 | 88.7 (10.2) | 90.4 (8.0) |  |  |
| | | Mean (SD) | Mean (SD) | $\beta$ (95% CI) | p value |
| **%NKG2A<sup>+</sup> CD56<sup>bright</sup> NK cells** |  |  |  |  |  |
| Exercise | 6 | 91.4 (4.7) | 92.1 (6.4) | 0.2 (-2.5; 2.9) | 0.85 |
| Control | 9 | 93.3 (3.2) | 94.0 (3.3) |  |  |
| **%KIR2DL1/DS1<sup>+</sup> CD56<sup>bright</sup> NK cells** |  |  |  |  |  |
| Exercise | 6 | 3.0 (2.6) | 2.2 (1.3) | 0.1 (-0.9; 1.1) | 0.84 |
| Control | 9 | 2.3 (1.2) | 1.7 (1.2) |  |  |
| **%KIR2DL2/DL3<sup>+</sup> CD56<sup>bright</sup> NK cells** |  |  |  |  |  |
| Exercise | 6 | 3.5 (1.9) | 3.1 (1.2) | -0.0 (-1.0; 0.9) | 0.94 |
| Control | 9 | 4.6 (1.3) | 3.7 (1.0)* |  |  |
| **%KIR3DL1<sup>+</sup> CD56<sup>bright</sup> NK cells** |  |  |  |  |  |
| Exercise | 6 | 2.2 (2.6) | 2.0 (2.0) | 0.3 (-0.8; 1.4) | 0.55 |
| Control | 9 | 2.9 (1.6) | 2.3 (1.6) |  |  |
| **%CD57<sup>+</sup> CD56<sup>bright</sup> NK cells** |  |  |  |  |  |
| Exercise | 6 | 1.4 (1.0) | 1.4 (1.2) | 0.3 (-0.8; 1.4) | 0.59 |
| Control | 9 | 0.8 (0.5) | 1.0 (0.6) |  |  |
| **%CD62L<sup>+</sup> CD56<sup>bright</sup> NK cells** |  |  |  |  |  |
| Exercise | 6 | 45.4 (14.2) | 34.9 (8.4) | -9.5 (-26.8; 7.7) | 0.25 |
| Control | 9 | 48.3 (21.1) | 44.2 (17.2) |  |  |
| **%CD69<sup>+</sup> CD56<sup>bright</sup> NK cells** |  |  |  |  |  |
| Exercise | 6 | 3.5 (1.4) | 3.8 (0.9) | 0.6 (-0.9; 2.0) | 0.41 |
| Control | 9 | 2.4 (1.0) | 2.7 (1.4) |  |  |
| **%DNAM-1<sup>+</sup> CD56<sup>bright</sup> NK cells** |  |  |  |  |  |
| Exercise | 6 | 94.7 (1.9) | 89.6 (7.1)* | 0.6 (-4.7; 5.9) | 0.82 |
| Control | 9 | 92.5 (6.5) | 84.9 (12.7)* |  |  |
| **%2B4<sup>+</sup> CD56<sup>bright</sup> NK cells** |  |  |  |  |  |
| Exercise | 4 | 87.8 (8.4) | 88.9 (2.6) | 2.2 (-2.6; 7.1) | 0.33 |
| Control | 9 | 82.3 (6.6) | 83.0 (6.5) |  |  |
| **%LIGHT<sup>+</sup> CD56<sup>bright</sup> NK cells** |  |  |  |  |  |
| Exercise | 4 | 2.1 (1.1) | 2.7 (1.6) | -0.0 (-2.0; 1.9) | 0.97 |
| Control | 9 | 2.5 (2.8) | 2.7 (1.3) |  |  |
| | | Mean (SD) | Mean (SD) | $\beta$ (95% CI) | p value |
| **%OX40<sup>+</sup> CD56<sup>bright</sup> NK cells** |  |  |  |  |  |
| Exercise | 4 | 0.2 (0.3) | 0.3 (0.3) | 0.1 (-0.7; 0.8) | 0.85 |
| Control | 9 | 0.5 (0.7) | 0.4 (0.8) |  |  |
| **%CD160<sup>+</sup> CD56<sup>bright</sup> NK cells** |  |  |  |  |  |
| Exercise | 4 | 6.7 (4.2) | 7.0 (2.5) | 2.0 (-0.9; 4.8) | 0.16 |
| Control | 9 | 6.6 (2.7) | 5.0 (2.3) |  |  |
| **%KLRG-1<sup>+</sup> CD56<sup>bright</sup> NK cells** |  |  |  |  |  |
| Exercise | 4 | 8.3 (4.9) | 9.6 (4.6) | 2.2 (-2.4; 6.8) | 0.32 |
| Control | 9 | 10.8 (9.4) | 9.6 (8.9) |  |  |
| **%SIGLEC7<sup>+</sup> CD56<sup>bright</sup> NK cells** |  |  |  |  |  |
| Exercise | 4 | 65.7 (8.7) | 62.6 (10.6) | 0.9 (-7.5; 9.4) | 0.81 |
| Control | 9 | 70.6 (5.3) | 66.2 (6.9)* |  |  |
| **%SIGLEC9<sup>+</sup> CD56<sup>bright</sup> NK cells** |  |  |  |  |  |
| Exercise | 4 | 0.1 (0.1) | 0.1 (0.1) | -0.1 (-0.3; 0.1) | 0.16 |
| Control | 9 | 0.1 (0.1) | 0.2 (0.1) |  |  |
| **%TIGIT<sup>+</sup> CD56<sup>bright</sup> NK cells** |  |  |  |  |  |
| Exercise | 4 | 23.5 (9.9) | 20.8 (4.9) | 3.3 (-2.3; 8.9) | 0.22 |
| Control | 9 | 20.5 (5.1) | 15.7 (5.6)* |  |  |
| **%CD96<sup>+</sup> CD56<sup>bright</sup> NK cells** |  |  |  |  |  |
| Exercise | 4 | 89.9 (4.2) | 90.4 (3.2) | -0.8 (-7.6; 6.0) | 0.79 |
| Control | 9 | 84.7 (6.1) | 88.5 (5.8) |  |  |
| **%TIM-3<sup>+</sup> CD56<sup>bright</sup> NK cells** |  |  |  |  |  |
| Exercise | 4 | 0.4 (0.5) | 0.2 (0.4) | -0.1 (-0.6; 0.4) | 0.77 |
| Control | 9 | 0.5 (0.5) | 0.4 (0.5) |  |  |
| **%PD-1<sup>+</sup> CD56<sup>bright</sup> NK cells** |  |  |  |  |  |
| Exercise | 4 | 0.9 (0.8) | 0.8 (0.9) | 0.3 (-0.3; 0.8) | 0.27 |
| Control | 9 | 0.9 (0.8) | 0.5 (0.4) |  |  |
| **%CD27<sup>+</sup> CD56<sup>bright</sup> NK cells** |  |  |  |  |  |
| Exercise | 4 | 53.1 (15.4) | 41.3 (21.2) | 2.7 (-9.2; 14.6) | 0.62 |
| Control | 9 | 62.0 (9.5) | 48.7 (12.4)* |  |  |

\*Significant within-group difference between T0 and T1 based on Wilcoxon signed rank (p<0.05).

**Supplementary Table 7.**
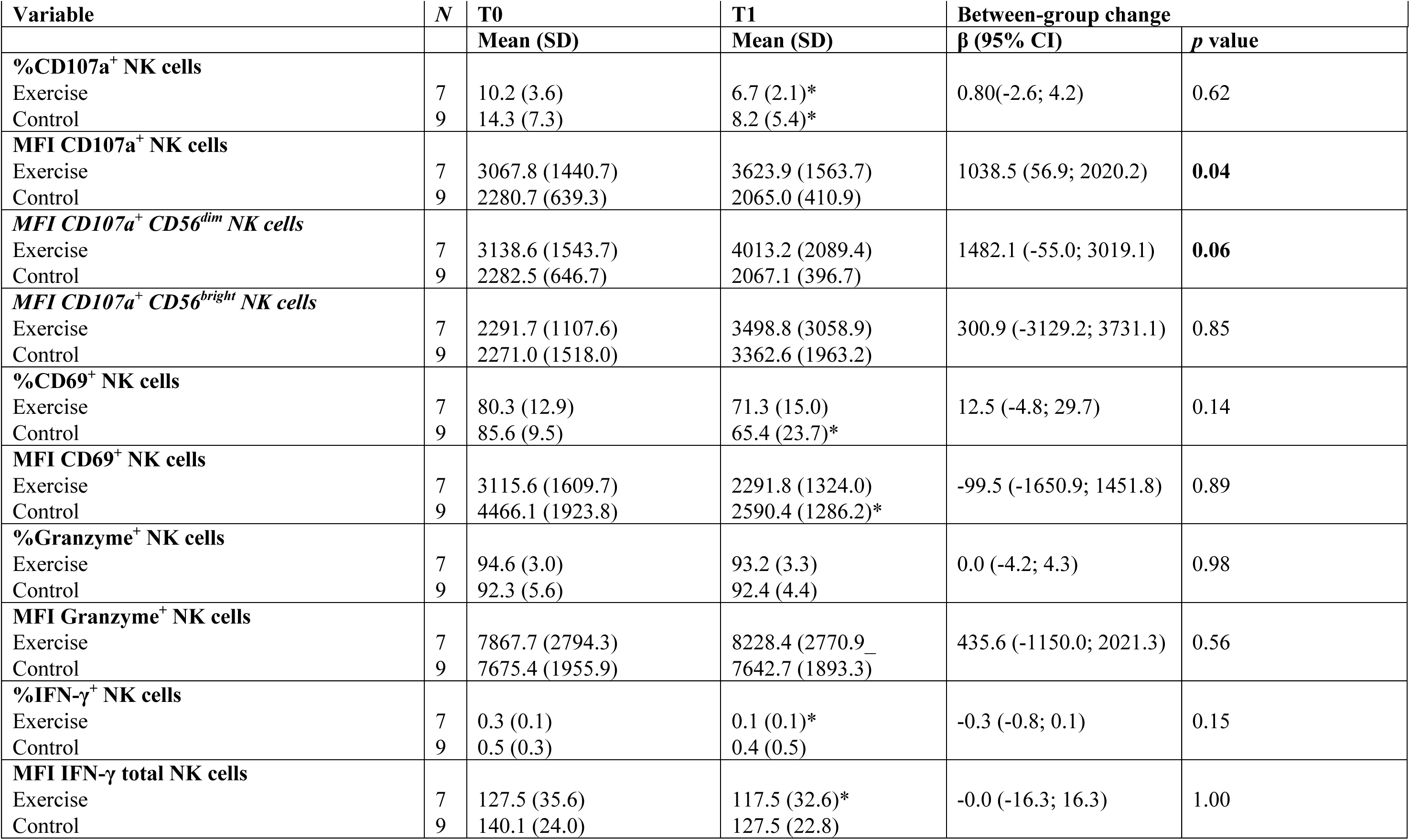
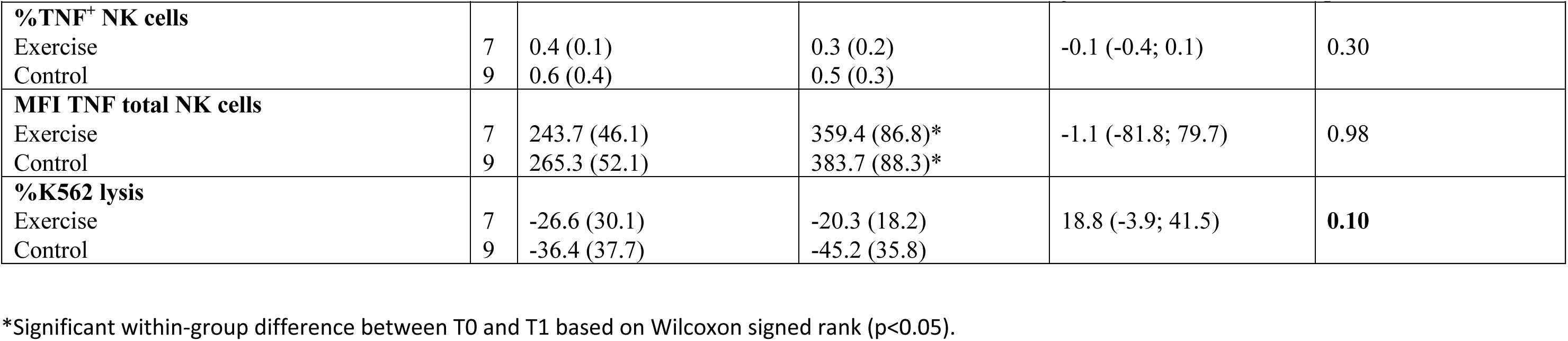
The percentage positive NK cells or the median fluorescence intensity (MFI) of the total or positive NK cell population after 4h co-culture with K562 tumour cells before (T0) and after (T1) 6 weeks of chemotherapy. The regression coefficients (β) indicate the between-group differences corrected for baseline values.

| Variable | N | T0 | T1 | Between-group change |  |
| --- | --- | --- | --- | --- | --- |
| | | Mean (SD) | Mean (SD) | $\beta$ (95% CI) | p value |
| **%CD107a<sup>+</sup> NK cells** |  |  |  |  |  |
| Exercise | 7 | 10.2 (3.6) | 6.7 (2.1)* | 0.80(-2.6; 4.2) | 0.62 |
| Control | 9 | 14.3 (7.3) | 8.2 (5.4)* |  |  |
| **MFI CD107a<sup>+</sup> NK cells** |  |  |  |  |  |
| Exercise | 7 | 3067.8 (1440.7) | 3623.9 (1563.7) | 1038.5 (56.9; 2020.2) | **0.04** |
| Control | 9 | 2280.7 (639.3) | 2065.0 (410.9) |  |  |
| **MFI CD107a<sup>+</sup> CD56<sup>dim</sup> NK cells** |  |  |  |  |  |
| Exercise | 7 | 3138.6 (1543.7) | 4013.2 (2089.4) | 1482.1 (-55.0; 3019.1) | **0.06** |
| Control | 9 | 2282.5 (646.7) | 2067.1 (396.7) |  |  |
| **MFI CD107a<sup>+</sup> CD56<sup>bright</sup> NK cells** |  |  |  |  |  |
| Exercise | 7 | 2291.7 (1107.6) | 3498.8 (3058.9) | 300.9 (-3129.2; 3731.1) | 0.85 |
| Control | 9 | 2271.0 (1518.0) | 3362.6 (1963.2) |  |  |
| **%CD69<sup>+</sup> NK cells** |  |  |  |  |  |
| Exercise | 7 | 80.3 (12.9) | 71.3 (15.0) | 12.5 (-4.8; 29.7) | 0.14 |
| Control | 9 | 85.6 (9.5) | 65.4 (23.7)* |  |  |
| **MFI CD69<sup>+</sup> NK cells** |  |  |  |  |  |
| Exercise | 7 | 3115.6 (1609.7) | 2291.8 (1324.0) | -99.5 (-1650.9; 1451.8) | 0.89 |
| Control | 9 | 4466.1 (1923.8) | 2590.4 (1286.2)* |  |  |
| **%Granzyme<sup>+</sup> NK cells** |  |  |  |  |  |
| Exercise | 7 | 94.6 (3.0) | 93.2 (3.3) | 0.0 (-4.2; 4.3) | 0.98 |
| Control | 9 | 92.3 (5.6) | 92.4 (4.4) |  |  |
| **MFI Granzyme<sup>+</sup> NK cells** |  |  |  |  |  |
| Exercise | 7 | 7867.7 (2794.3) | 8228.4 (2770.9) | 435.6 (-1150.0; 2021.3) | 0.56 |
| Control | 9 | 7675.4 (1955.9) | 7642.7 (1893.3) |  |  |
| **%IFN-γ<sup>+</sup> NK cells** |  |  |  |  |  |
| Exercise | 7 | 0.3 (0.1) | 0.1 (0.1)* | -0.3 (-0.8; 0.1) | 0.15 |
| Control | 9 | 0.5 (0.3) | 0.4 (0.5) |  |  |
| **MFI IFN-γ total NK cells** |  |  |  |  |  |
| Exercise | 7 | 127.5 (35.6) | 117.5 (32.6)* | -0.0 (-16.3; 16.3) | 1.00 |
| Control | 9 | 140.1 (24.0) | 127.5 (22.8) |  |  |
| | | Mean (SD) | Mean (SD) | $\beta$ (95% CI) | p value |
| **%TNF<sup>+</sup> NK cells** |  |  |  |  |  |
| Exercise | 7 | 0.4 (0.1) | 0.3 (0.2) | -0.1 (-0.4; 0.1) | 0.30 |
| Control | 9 | 0.6 (0.4) | 0.5 (0.3) |  |  |
| **MFI TNF total NK cells** |  |  |  |  |  |
| Exercise | 7 | 243.7 (46.1) | 359.4 (86.8)* | -1.1 (-81.8; 79.7) | 0.98 |
| Control | 9 | 265.3 (52.1) | 383.7 (88.3)* |  |  |
| **%K562 lysis** |  |  |  |  |  |
| Exercise | 7 | -26.6 (30.1) | -20.3 (18.2) | 18.8 (-3.9; 41.5) | **0.10** |
| Control | 9 | -36.4 (37.7) | -45.2 (35.8) |  |  |

\*Significant within-group difference between T0 and T1 based on Wilcoxon signed rank (p<0.05).

**Supplementary Table 8.** Percentage positive NK cells or the median fluorescence intensity (MFI) of the total or positive NK cell population after overnight stimulation with IL2/IL15 and subsequent 4h co-culture with K562 tumour cells before (T0) and after (T1) six weeks of chemotherapy.

| Variable | N | T0 | T1 | Between-group change |  |
| --- | --- | --- | --- | --- | --- |
| | | Mean (SD) | Mean (SD) | $\beta$ (95% CI) | p value |
| **%CD107a<sup>+</sup> NK cells IL2/IL15** |  |  |  |  |  |
| Exercise | 7 | 48.3 (4.8) | 44.6 (6.4) | -4.1 (-10.5; 2.3) | 0.19 |
| Control | 9 | 51.8 (5.6) | 50.4 (5.5) |  |  |
| **MFI CD107a<sup>+</sup> NK cells IL2/IL15** |  |  |  |  |  |
| Exercise | 7 | 3327.5 (1003.9) | 3095.7 (892.1) | 198.2 (-465.6; 862.0) | 0.53 |
| Control | 9 | 3288.9 (799.1) | 2872.3 (774.7) |  |  |
| **%CD69<sup>+</sup> NK cells IL2/IL15** |  |  |  |  |  |
| Exercise | 7 | 99.6 (0.2) | 99.2 (0.4)* | -0.10 (-0.74; 0.54) | 0.75 |
| Control | 9 | 99.5 (0.3) | 99.3 (0.7) |  |  |
| **MFI CD69<sup>+</sup> NK cells IL2/IL15** |  |  |  |  |  |
| Exercise | 7 | 9025.1 (1881.8) | 6295.3 (1155.0)* | -524.5 (-2495.8; 1446.8) | 0.58 |
| Control | 9 | 9744.8 (993.4) | 7053.9 (2089.0)* |  |  |
| **%Granzyme<sup>+</sup> NK cells IL2/IL15** |  |  |  |  |  |
| Exercise | 7 | 99.5 (0.6) | 99.4 (0.4) | 0.0 (-0.4; 0.5) | 0.84 |
| Control | 9 | 99.5 (0.7) | 99.3 (0.5) |  |  |
| **MFI Granzyme<sup>+</sup> NK cells IL2/IL15** |  |  |  |  |  |
| Exercise | 7 | 15093.0 (3789.0) | 16267.2 (3126.3) | -1396.1 (-5255.0; 2462.7) | 0.45 |
| Control | 9 | 18373.6 (5898.5) | 19340.4 (4781.6) |  |  |
| **%IFN-γ<sup>+</sup> NK cells IL2/IL15** |  |  |  |  |  |
| Exercise | 7 | 8.0 (4.0) | 6.0 (2.8) | -2.1 (-6.7; 2.6) | 0.35 |
| Control | 9 | 13.9 (7.4) | 9.3 (4.6) |  |  |
| **MFI IFN-γ total NK cells IL2/IL15** |  |  |  |  |  |
| Exercise | 7 | 316.0 (116.5) | 252.6 (96.1)* | -33.6 (-128.7; 61.5) | 0.46 |
| Control | 9 | 407.1 (143.2) | 317.0 (86.8) |  |  |
| **%TNF<sup>+</sup> NK cells IL2/IL15** |  |  |  |  |  |
| Exercise | 7 | 6.6 (3.0) | 6.0 (2.6) | -2.5 (-6.3; 1.2) | 0.17 |
| Control | 9 | 7.2 (2.4) | 9.0 (4.7) |  |  |
| **MFI TNF total NK cells IL2/IL15** |  |  |  |  |  |
| Exercise | 7 | 313.1 (67.7) | 414.9 (109.1)* | -13.4 (-114.7; 87.9) | 0.78 |
| Control | 9 | 328.0 (68.5) | 439.3 (96.8)* |  |  |
| | | Mean (SD) | Mean (SD) | $\beta$ (95% CI) | p value |
| %K562 lysis IL2/IL15 |  |  |  |  |  |
| Exercise | 7 | 43.5 (32.6) | 15.1 (38.3)* | -0.5 (-28.9; 28.0) | 0.97 |
| Control | 9 | 39.9 (29.9) | 13.3 (26.1)* |  |  |

\*Significant within-group difference between T0 and T1 based on Wilcoxon signed rank ( $p < 0.05$ ).

**Supplementary Figure 1.**
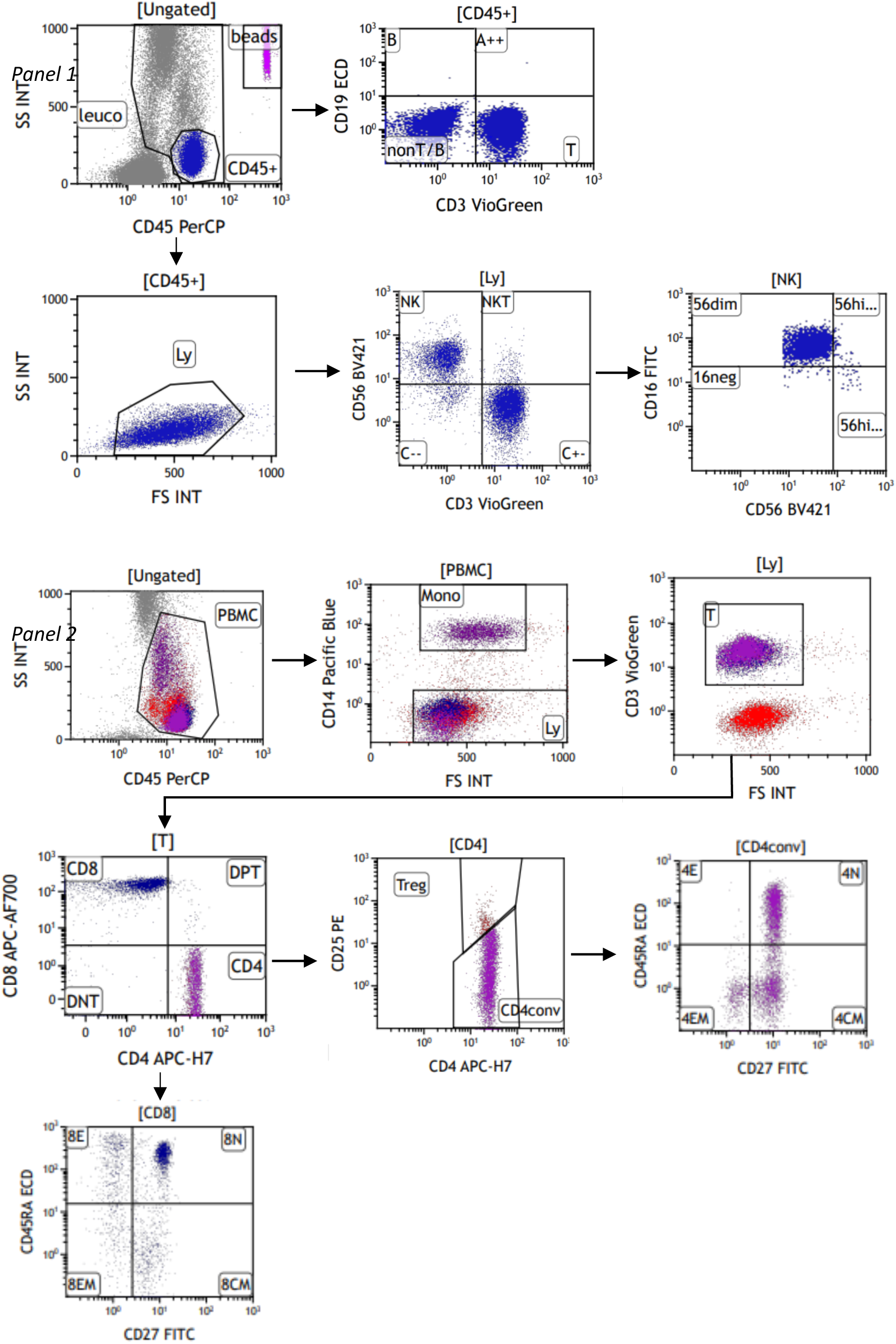
Gating strategy immune cell profiling.

**Supplementary Figure 2.**
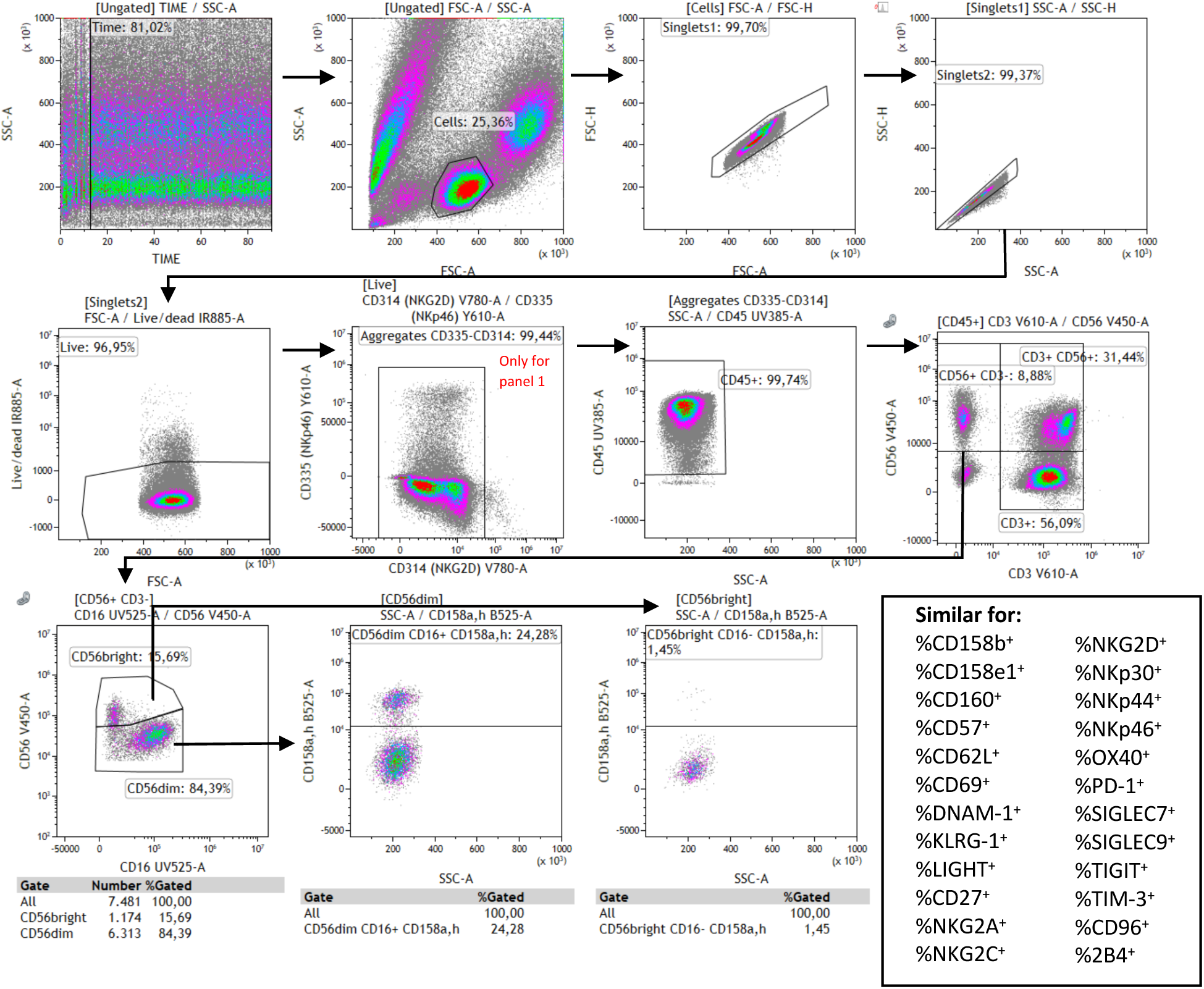
Gating strategy NK cell phenotype.

**Supplementary Figure 3.**
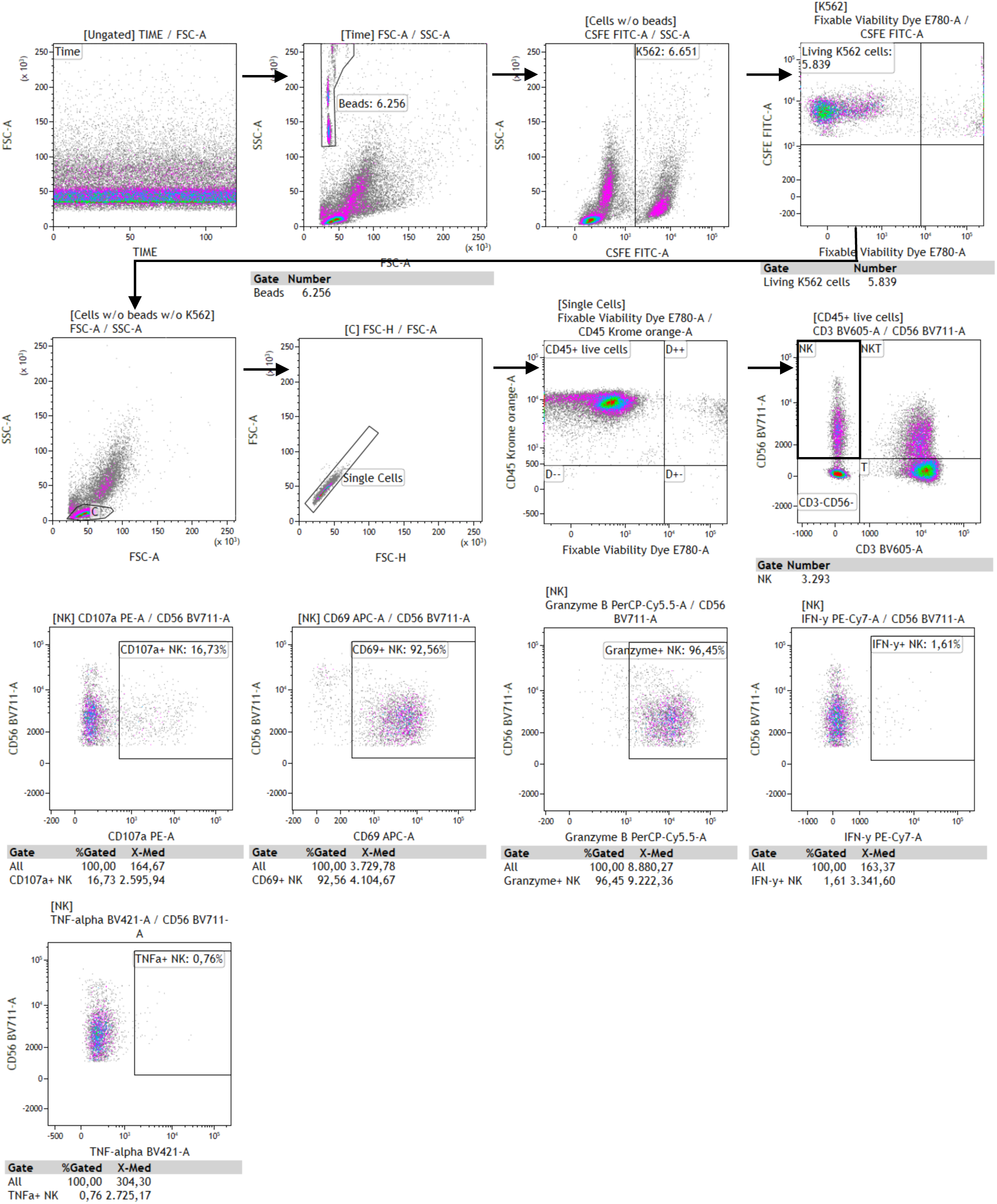
Gating strategy NK cell function.

**Supplementary Figure 4.**
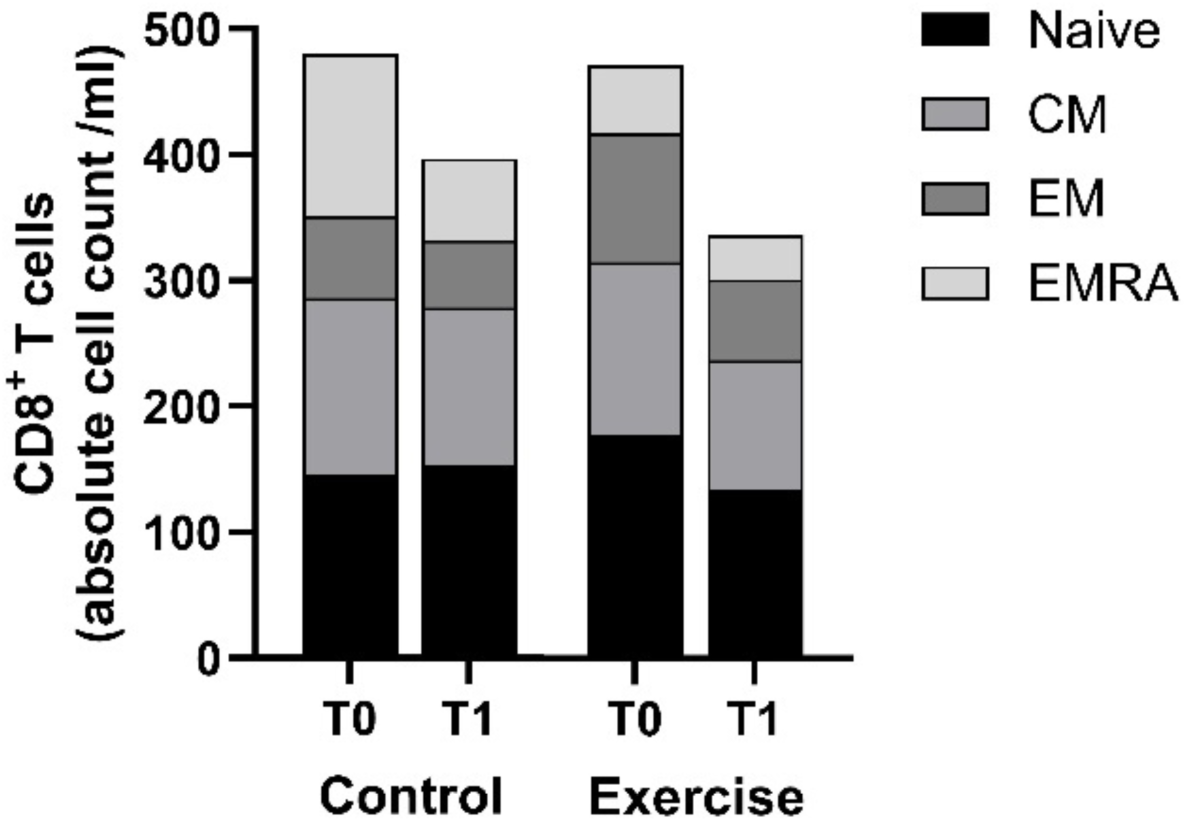
Absolute counts of CD8^+^ T cell subsets. Absolute counts of the CD8^+^ T cell subsets per ml peripheral blood of patients with breast cancer from the exercise intervention (n=7) and control group (n=9) before (T0) and after (T1) six weeks of neoadjuvant chemotherapy. Linear regression analysis showed no significant between-group differences in the absolute cell counts of the CD8^+^ T cell subsets. Data is presented as mean values. Abbreviations: CM, central memory; EM, effector memory; EMRA, effector memory CD45RA^+^.

**Supplementary Figure 5.**
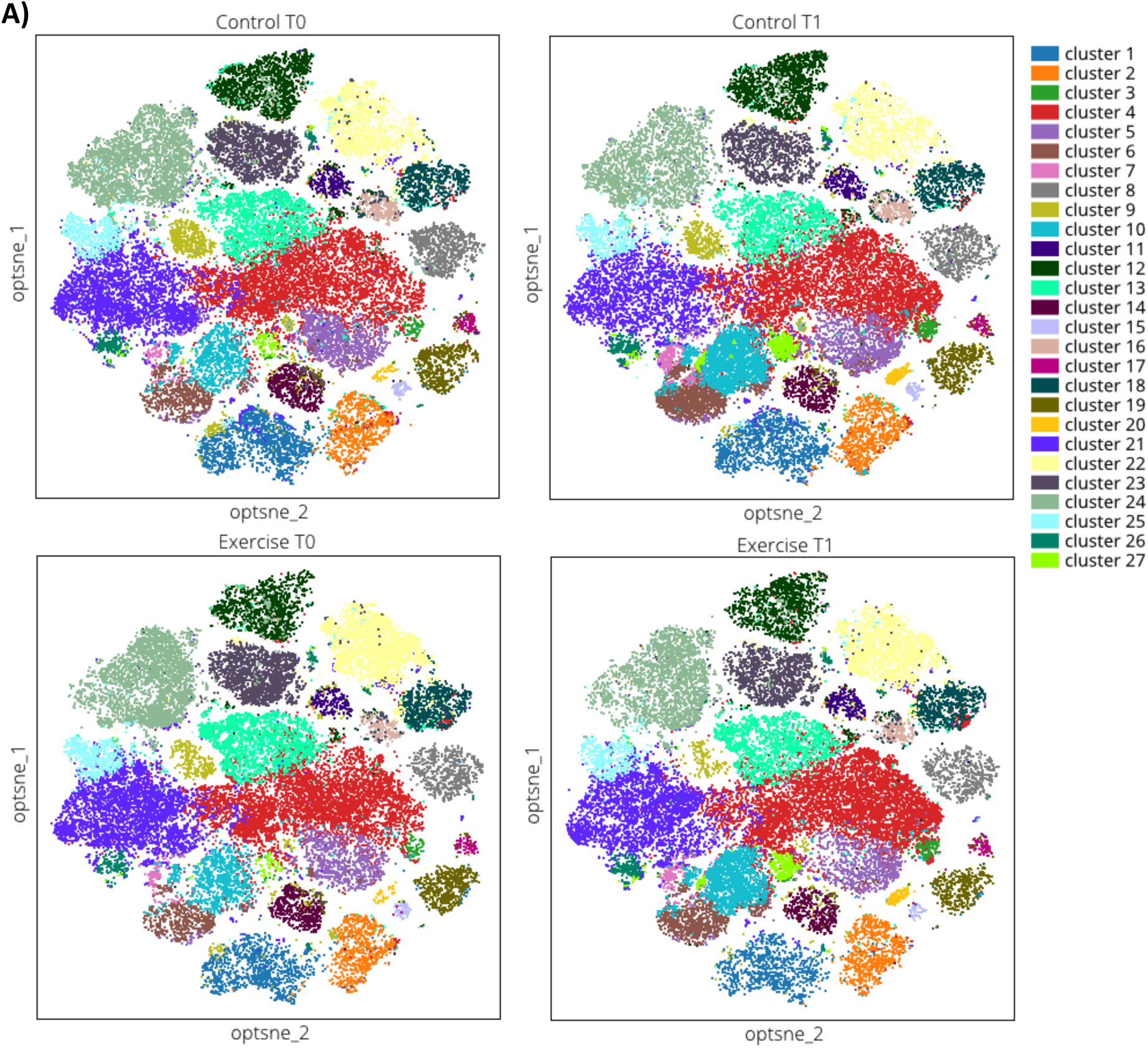
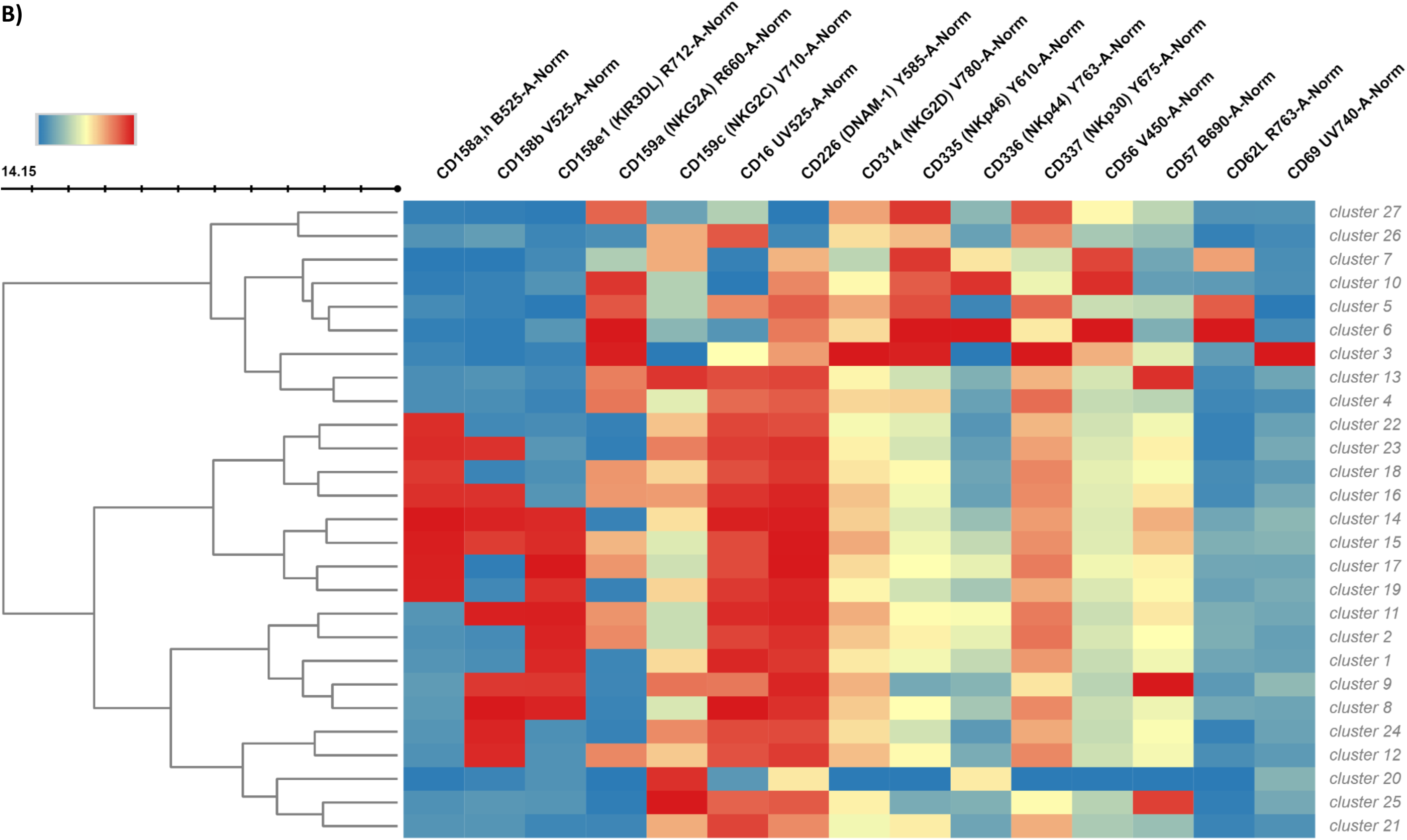
Cluster analysis of NK cell phenotype. **A)** Cluster analysis of NK cells based on the expression of activating and inhibitory receptors in the exercise intervention (n=6) and control group (n=9) before (T0) and after (T1) six weeks of neoadjuvant chemotherapy. **B)** (next page) Heatmap showing the expression of each activating or inhibitory marker within each NK cell cluster. Red = high expression, blue = low expression.

**Supplementary Figure 6.**
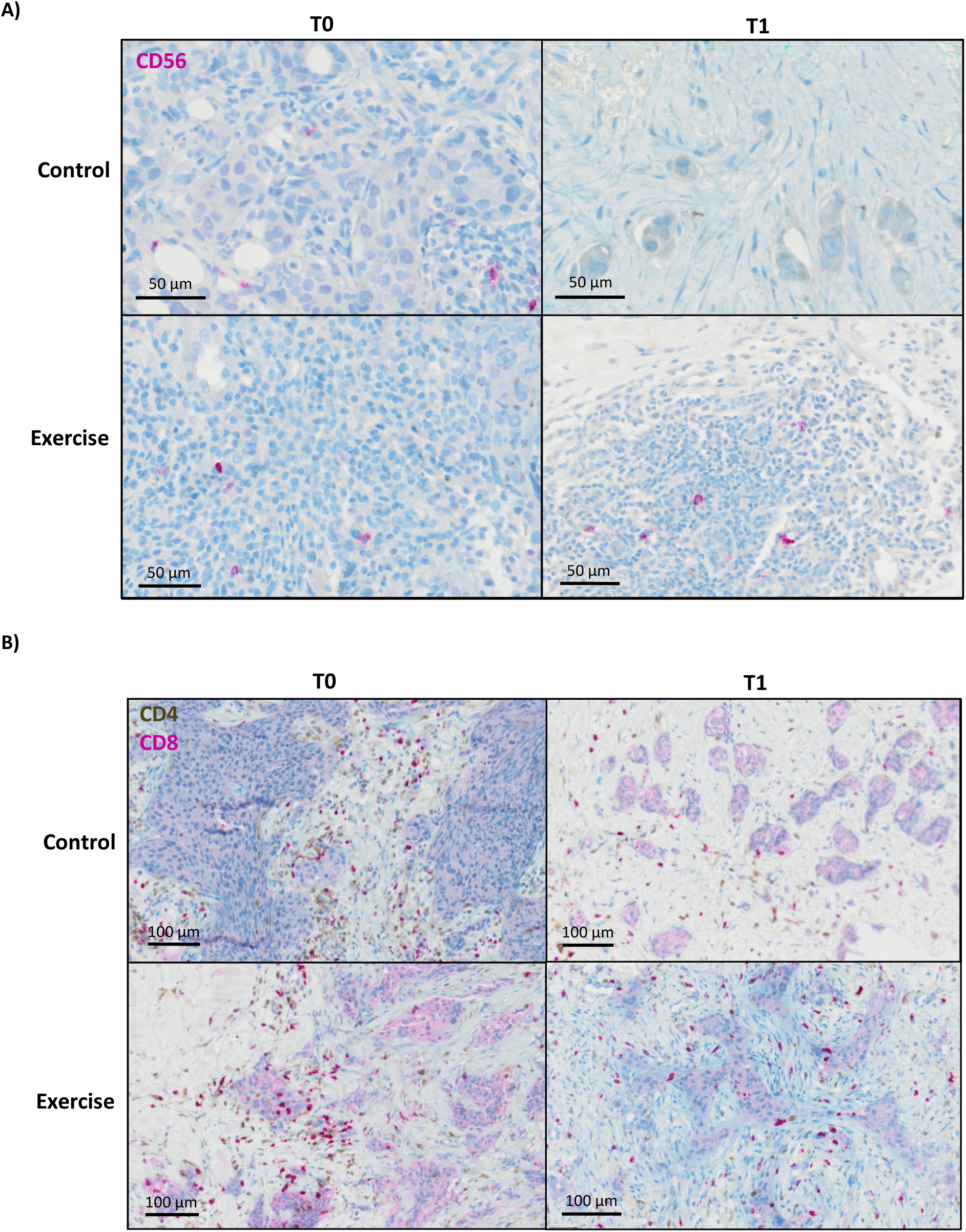
Tumour infiltrating lymphocytes. The infiltration of **A)** CD56^+^ cells (red) and **B)** CD4^+^ (brown) and CD8^+^ (red) cells in tissue sections from biopsies of illustrative patients of the control group and the exercise group at T0 (baseline) and T1 (after six weeks of neoadjuvant chemotherapy).

## Notes

### Competing Interest Statement

The authors have declared no competing interest.

### Clinical Trial

NCT04704856

### Author Declarations

Ethics committee of Amsterdam University Medical Centres gave ethical approval for this work

